# Diagnosis of SARS-Cov-2 infection using specimens other than naso- and oropharyngeal swabs: a systematic review and meta-analysis

**DOI:** 10.1101/2021.01.19.21250094

**Authors:** Vânia M. Moreira, Paulo Mascarenhas, Vanessa Machado, João Botelho, José João Mendes, Nuno Taveira, M. Gabriela Almeida

## Abstract

**Background:** The rapid and accurate testing of SARS-CoV-2 infection is still crucial to mitigate, and eventually halt, the spread of this disease. Currently, nasopharyngeal swab (NPS) and oropharyngeal swab (OPS) are the recommended standard sampling, yet, with some limitations. Several specimens that are easier to collect are being tested as alternatives to nasal/throat swabs in nucleic acid assays for SARS-CoV-2 detection. This study aims to critically appraise and compare the clinical performance of RT-PCR tests using oral saliva, deep-throat saliva/ posterior oropharyngeal saliva (DTS/POS), sputum, urine, feces, and tears/conjunctival swab [CS]) against standard specimens (NPS, OPS, or a combination of both).

**Methods:** In this systematic review and meta-analysis, five databases (PubMed, Scopus, Web of Science, ClinicalTrial.gov and NIPH Clinical Trial) were searched up to the 30^th^ of December 2020. Case-control and cohort studies on the detection of SARS-CoV-2 were included. Methodological quality was assessed through the Quality Assessment of Diagnostic Accuracy Studies 2 (QUADAS 2).

**Findings:** We identified 3022 entries, 33 of which (1.1%) met all required criteria and were included for the quantitative data analysis. Saliva presented the higher accuracy, 92.1% (95% CI: 70.0-98.3), with an estimated sensitivity of 83.9% (95% CI: 77.4-88.8) and specificity of 96.4% (95% CI: 89.5-98.8). DTS/POS samples had an overall accuracy of 79.7% (95% CI: 43.3-95.3), with an estimated sensitivity of 90.1% (95% CI: 83.3-96.9) and specificity of 63.1% (95% CI: 36.8-89.3). Remaining index specimens presented uncertainty given the lack of studies available.

**Interpretation:** Our meta-analysis shows that saliva samples from oral region provide a high sensitivity and specificity, being the best candidate as an alternative specimen to NPS/OPS for COVID-19 detection, with suitable protocols for swab-free sample collection to be determined and validated in the future. The distinction between oral and extra-oral salivary samples will be crucial since DTS/POS samples may induce a higher rate of false positives. Urine, feces, tears/CS and sputum seem unreliable for diagnosis. Saliva testing may increase testing capacity, ultimately promoting the implementation of truly deployable COVID-19 tests, which could either work at the point-of-care (e.g. hospitals, clinics) or outbreak control spots (e.g. schools, airports, and nursing homes).

**Funding:** Nothing to declare.

**Research in context:** *Evidence before this study:* The lack of systematized data on the accuracy performance of alternative specimens for the detection of SARS-CoV-2 (against the standard NPS/OPS). The ever-growing number of studies available, made this updated systematic review timely and of the utmost importance

*Added value of this study:* Our meta-analysis shows that saliva samples from the oral region provide a high sensitivity and specificity, being the best candidate as an alternative specimen to NPS/OPS for COVID-19 detection, with suitable protocols for swab-free sample collection to be determined and validated in the future. The distinction between oral and extra-oral salivary samples will be crucial since DTS/POS samples may induce a higher rate of false positives.

*Implications of all the available evidence:* Saliva samples simply taken from the oral cavity are promising alternatives to the currently used nasal/throat swabs. Saliva specimens can be self-collected, mitigate the discomfort caused by sampling, reduce the transmission risk and increase testing capacity. Therefore, the validation of this alternative specimen will promote the implementation of truly deployable rapid tests for SARS-CoV-2 detection at the point-of-care or outbreak spots.

## Introduction

The COVID-19 outbreak was labeled a pandemic by the World Health Organization (WHO) on March 11th 2020. Since then, COVID-19 has been rapidly spreading around the globe. By the end of 2020, the number of deaths had totaled more than 1.7 M, and 80 M people had tested positive for SARS-CoV-2 worldwide, though the actual numbers are expected to be much higher ^1^. One of the greatest challenges of SARS-CoV-2 is its high transmissibility rate, drastically increasing the number of infected people in a short amount of time ^2,3^. Timely and reliable diagnosis is, thus, vital in preventing the spread of COVID-19, and a huge effort has been made to test as many people at risk as possible, regardless of being symptomatic or not. On the one hand, test results allow physicians to promptly treat and isolate viral carriers, which otherwise would result in further transmissions and even more devastating outbreaks. Additionally, testing ensures a better understanding of the disease’s progression and public health management as well as the pandemic’s epidemiology ^4^. Diagnosis of SARS-Cov-2 infection can be done in three different ways. Direct diagnostic assays target the viral RNA genome (NUC assays) or a viral antigen (antigen assays), which typically is a viral surface protein. Indirect antibody assays assess the human immune response to the coronavirus infection ^4,5^. The amplification of viral RNA using Real-Time Reverse-Transcription Polymerase Chain Reaction (RT-PCR) technology is the gold standard test to confirm SRAS-cov-2 infection. Specimens are collected from the upper respiratory tract (URT) such as nasopharyngeal swabs (NPS) and/or oropharyngeal swabs (OPS) since the viral load tends to be higher therein, thus improving the sensitivity and reliability of the results ^5–7^. However, this procedure requires training and specific cautions, especially when dealing with elderly people or children ^3,8^, and with patients that have had recent nasal trauma or have a deviated nasal septum, among other complications ^9^. In addition, it can cause discomfort to patients, and may pose a high risk of transmission, putting greater strain on resources (such as protective equipment) and on professionals ^5,6,10^.

The urgent demand of test kits for decentralised detection of SARS-CoV-2 infections has fueled a new frontier of diagnostic innovation. Initially, miniaturized systems for nucleic acid tests based in PCR technology were used. Currently, new commercial *in vitro* diagnostic medical devices (IVDs) are being used in antigen testing at the point-of-care or even in laboratory settings, such as the rapid tests provided by Abbott (Panbio COVID-19 Ag), RapiGen (Biocredit COVID19), Liming Bio-Products (StrongStep COVID-19), Savant Biotechnology (Huaketai New Coronavirus), and Bioeasy Biotechnology (Diagnostic Kit for 2019-nCoV Ag Test), among others. However, these tests are validated for URT swabs only ^4^.

Aiming at simplifying the sample collection procedure, so that the average person could perform self-sampling, alternative specimens have been tested for the detection of SARS-CoV-2, namely sputum (or bronchoalveolar lavage), saliva, tears/conjunctival swab (CS), feces, rectal swab, urine, breast milk, and semen ^11–42^. To the best of our knowledge, up to now, just one protocol for saliva testing, the SalivaDirect, has been approved by a recognized public health authority, the FDA ^43^. However, the accuracy of saliva-based tests for clinical use is still controversial. A preliminary meta-analysis published in August 2020 revealed that the sensitivity of saliva tests is promising (91%), although lower than nasal swabs based assays (98%) ^3^. The lack of data on specificity did not allow a statistically significant analysis of this parameter and therefore, on the tests’ accuracy. Probably, the main problem resided in the high variety and heterogeneity of studies (and results) for each specimen^3^.

Considering the ever-growing publication of scientific articles comparing alternative specimens for SARS-Cov-2 infection diagnosis, a more comprehensive and systematized review of the currently available literature providing meta-analytical estimates would be timely and of the utmost importance.

In this way, we aim at contributing to clarify whether specimens other than the conventional nasal/throat swab specimens can be used to diagnose and manage SARS-Cov-2 infection. Therefore, we have systematically appraised and compared the overall accuracy of nucleic acid assays run with index specimens (saliva, deep-throat saliva/posterior oral samples [DTS/POS], sputum, urine, feces, and tears/CS), against standard NPS/OPS based test results.

## 2. Materials and Methods

### 2.1. Protocol

This systematic review was submitted to PROSPERO (ID: CRD42021223894) and used the Preferred Reporting Items for Systematic reviews and Meta-analysis (PRISMA) guidelines ^44^. The PRISMA checklist is available as a Supplemental information file (appendix S1, pp 2-3).

### 2.2. Focused question and eligibility criteria

The following PECO question was set: “Are physiological specimens collected without invasive swabs as accurate as the NPS/OPS specimens in the detection of SARS-CoV-2 infection by nucleic acid assays?”.

Studies were deemed eligible as per the following criteria:

a. Observational studies (i.e. cross-sectional, case-control or cohort study types)
b. Use of RT-PCR to detect the presence of SARS-CoV-2 in matched samples;
c. Report SARS-CoV-2 positive and negative test results, and/or cycle threshold (CT) from index alternative specimens (saliva, DTS/POS, sputum, urine, feces, or tears/CS) evaluated against NPS and/or OPS;
d. Studies with confirmed or suspected cases of SARS-CoV-2 infection.

We also included CT (number of cycles needed to amplify viral RNA to reach a detectable level) as a complementary measure of sensitivity in matched samples.

Saliva samples refer to samples collected from the oral region (i.e., circumscribed to the oral cavity) while DTS/POS refers to salivary samples mixed with pharyngeal secretions. Sputum refers to primarily lower respiratory tract mucous mixed with pharyngeal and salivary secretions.

### 2.3. Search strategy and study selection

Search strategies were carried out in different databases (PubMed, Scopus, Web of Science, ClinicalTrial.gov and NIPH Clinical Trial) until December 4th, 2020. The search was repeated on 30th of December 2020 to confirm newly potential eligible studies.

We used the following search syntax: (COVID-19 OR COVID19 OR n-CoV19 OR SARS-CoV-2 OR SARS-CoV2) AND (Diagnosis OR Diagnostic OR Test OR Detection) OR (Saliva OR Salivary OR “Oral fluid” OR Sputum OR Expectoration OR Gob OR Tears OR Conjunctival OR Stool OR Feces OR Fecal OR Urine). No restrictions on the year of publication nor on language were made. We used Mendeley reference manager to organize records and remove duplicates. The study selection was assessed independently by two investigators (V.M.M. and P.M.), and by screening the titles and abstracts of retrieved studies. Articles selected at this point were further appraised by full text reading.

Inter-examiner reliability after full-text assessment was computed through Cohen’s kappa statistics, and any disagreements were resolved by discussion with a third author (M.G.A.).

### 2.4. Data extraction process and data items

Two authors (V.M.M. and P.M.) independently retrieved and reviewed the following data (if available) from all included studies: year of publication, first author, location, design, population size, mean age, gender ratio, mean days after symptoms onset, specimens and methods used; and the following test outcomes: number of total, positives, negatives, and CTs.

### 2.5. Risk of bias assessment

The methodological quality of the included studies was evaluated independently by two authors (V.M.M. and P.M.), using the Quality Assessment of Diagnostic Accuracy Studies 2 (QUADAS-2) tool ^45^, with any discordant rating resolved by a third author (M.G.A.). This instrument judges the risk of bias (RoB) and accessibility from diagnostic accuracy studies. QUADAS-2 contains four key domains (patient selection, index test, reference standard, and flow and timing) and each domain is rated as low, high, and unclear RoB. The *robvis* tool was used to generate all the RoB plots ^46^. If a study failed to provide enough information, the domain was classified as “No information”.

### 2.6. Quantitative analyses

We used MetaDTA ^47^ to examine the overall SARS-CoV-2 detection test accuracy and perform subgroup sensitivity analysis for the selected index specimens. In MetaDTA, the bivariate random-effects model meta-analyses pooled estimates for sensitivity and specificity together. This approach accounts for potential threshold effects and covariance between sensitivity and specificity. However, because these two parameters depend on many other factors, accuracy heterogeneity is expected to be high and problematic to estimate ^48^. Diagnostic Odds Ratios (dOR) were directly obtained from the sensitivity and specificity logit estimates. Furthermore, the summary Receiver Operating Characteristic (sROC) plot was rendered using parameters estimated from the bivariate model through the equivalence equations of Harbord et al ^49^. CTs random effects meta-analysis, and all meta-regressions to identify potential sources of heterogeneity or confounding within or between the evaluated index specimens meta-analysis were performed with OpenMeta-analyst ^50^. The influence of the specific time of sampling and the disease stage on the accuracy rate of the test were planned to be assessed through meta-regression.

## 3 Results

Electronic searches revealed a total of 3022 entries (1406 articles from PubMed, 522 from Web of Science and 1094 from Scopus). After removing replicates, 1560 articles were judged against the eligibility criteria, and 1415 were excluded after title and/or abstract review. Out of the 145 subjected to full paper review, 112 articles were excluded (appendix S2, pp 4-11). As a result, a final of 33 studies met all the required criteria and were included for the quantitative data analysis (Figure 1). Inter-examiner agreement was considered as almost perfect agreement (k= 0.907, 95% CI: 0.828-0.987).

**Figure 1.**
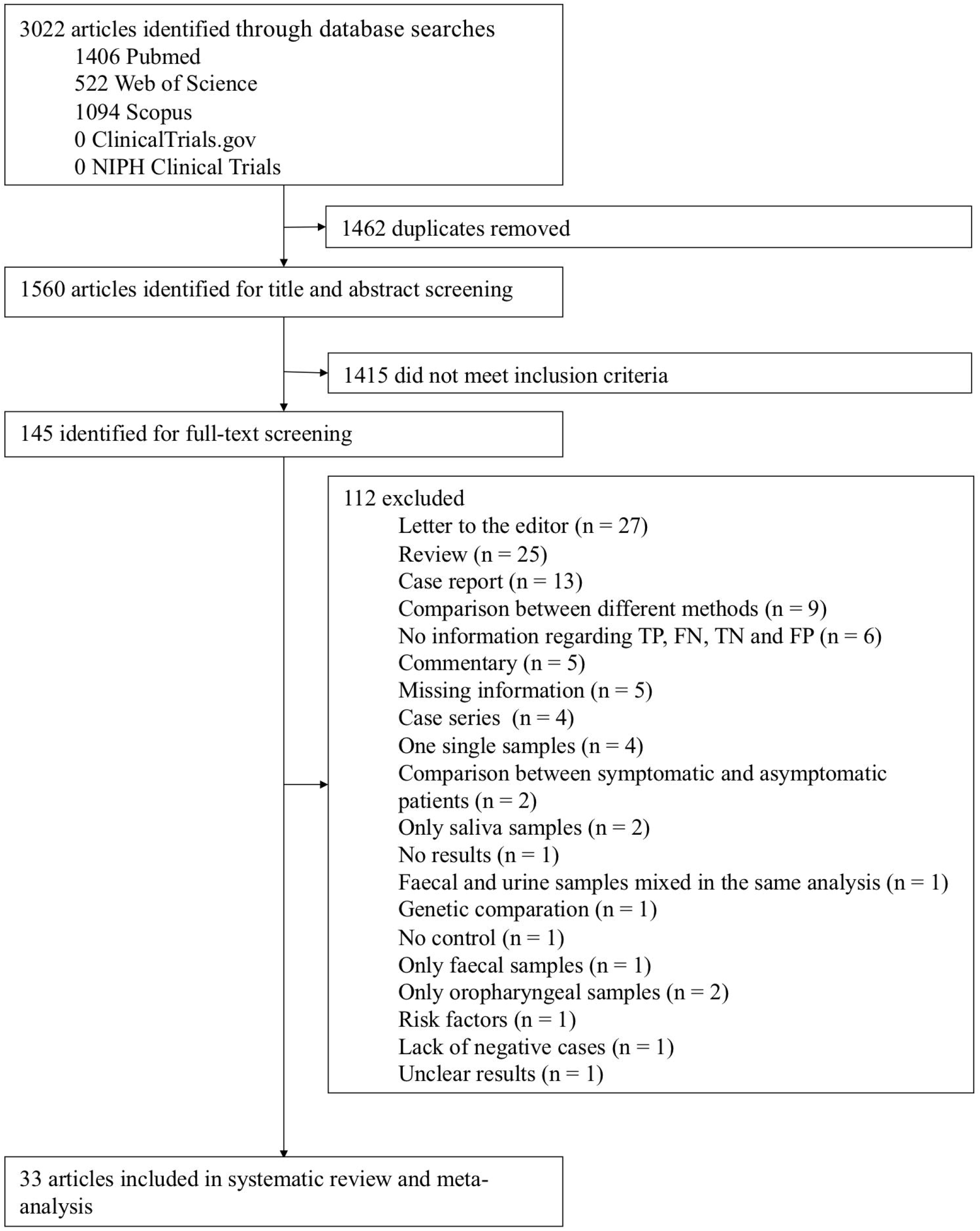
Flowchart of the included studies.

### 3.1. Characterization of the studies

All studies used a PCR-based method using different targets (E, N, ORFab1, or RdRP) and compared NPS and/or OPS samples with index specimens (sputum, saliva, DTS/POS, feces, tears/CS, and urine). Twelve articles did not provide information about the control used ^25,27–30,34–37,41,42,51^, yet the majority used RNase P. The main characteristics of the included studies are listed in Table 1.

**Table 1.**
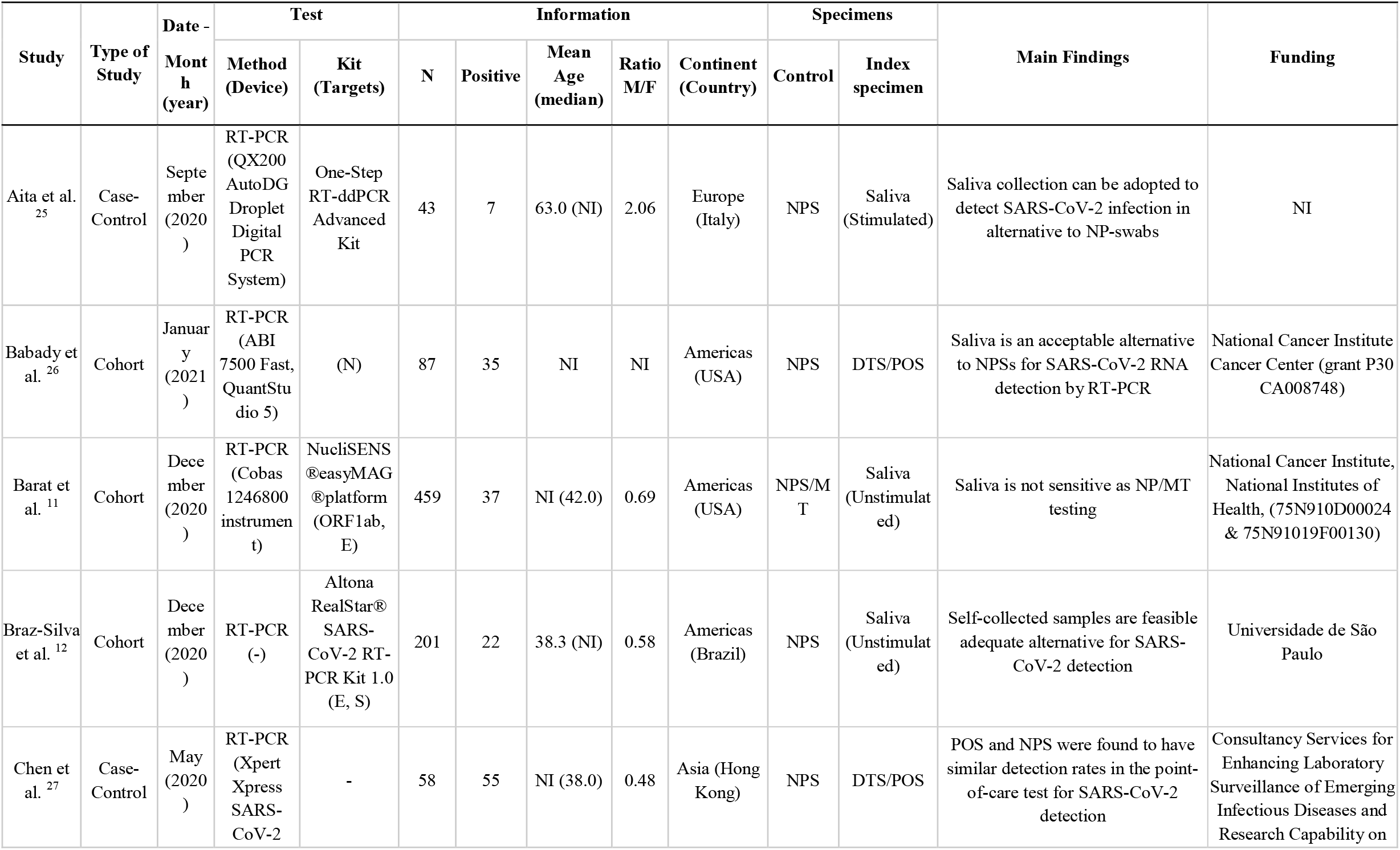
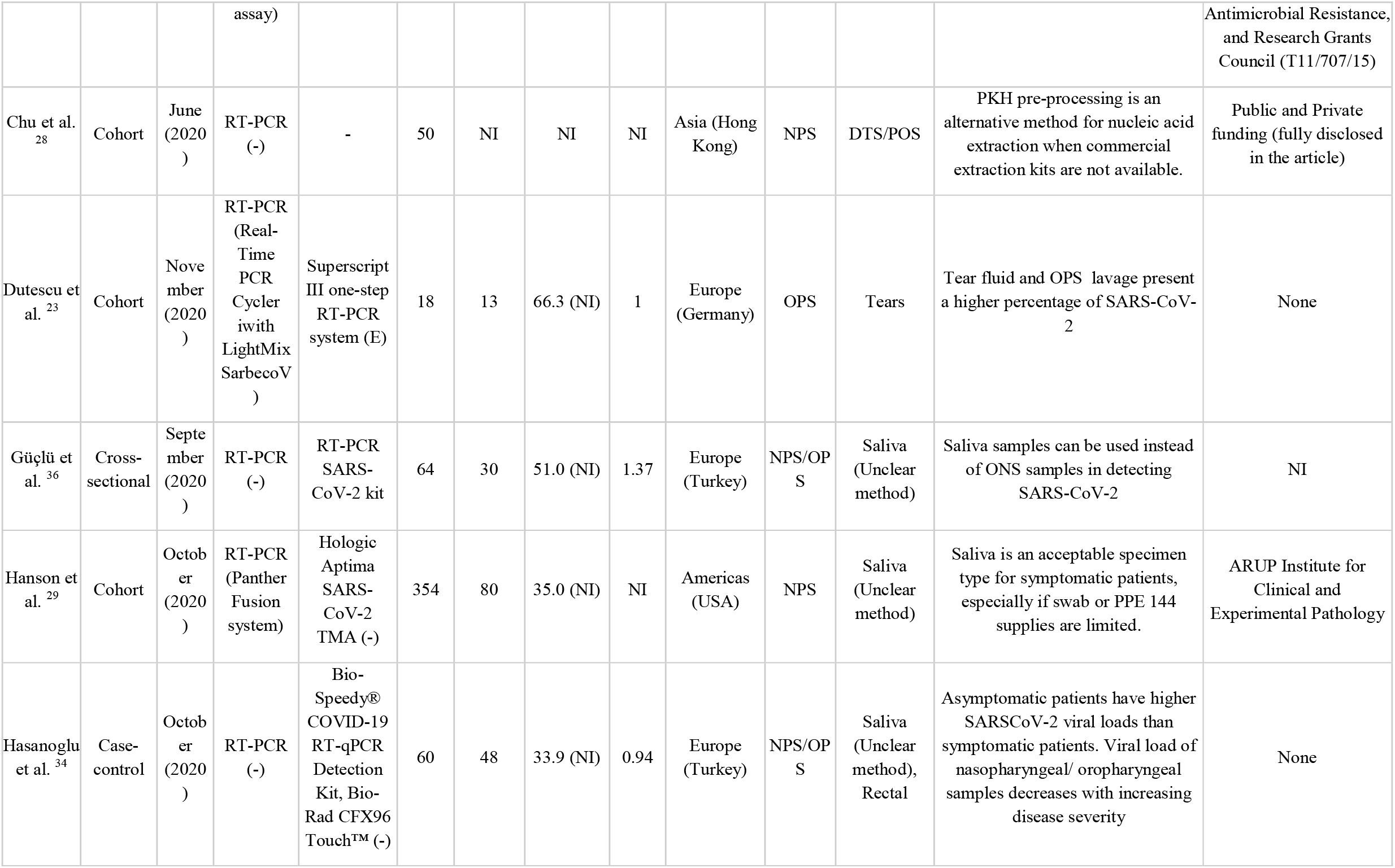
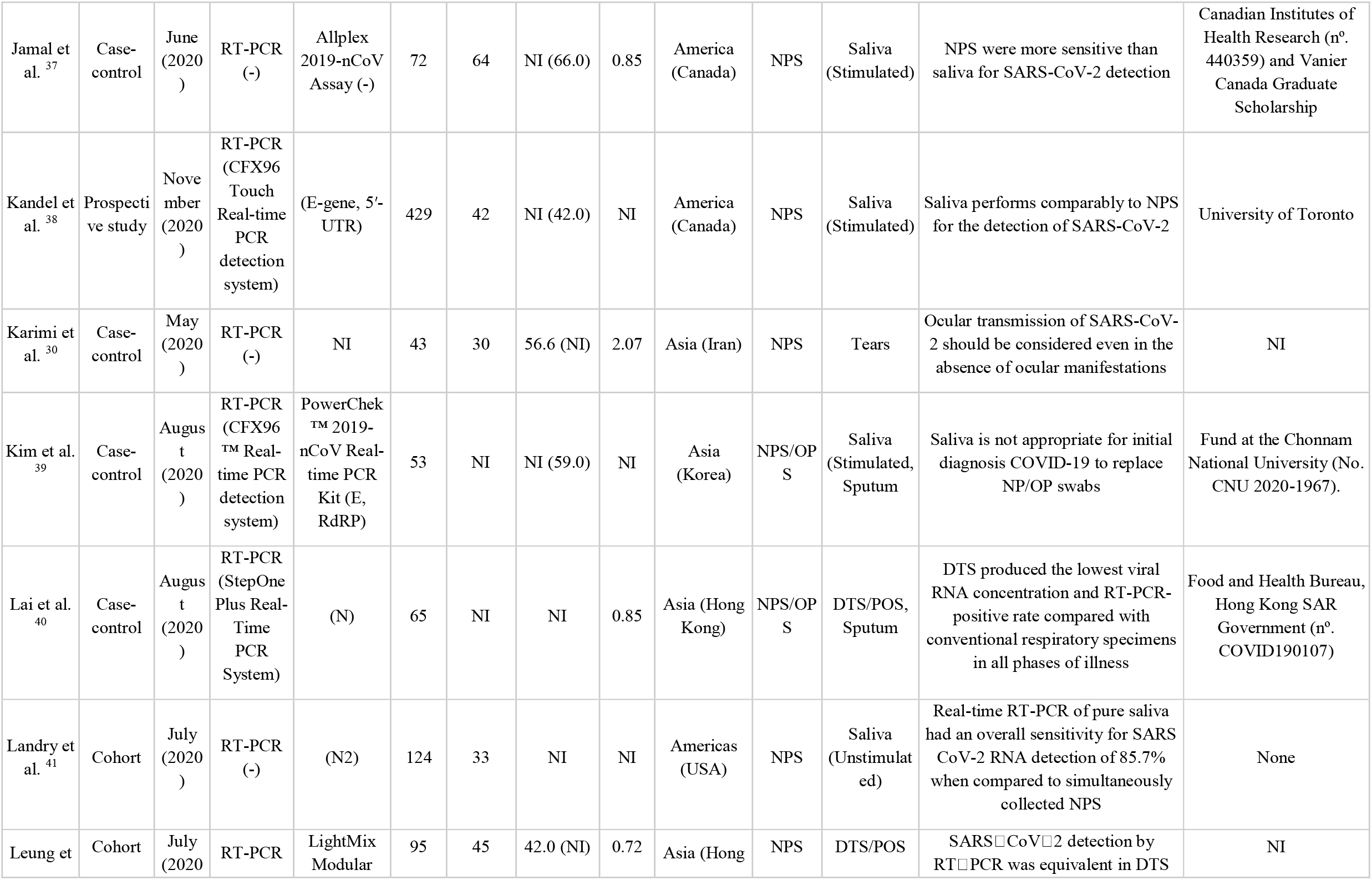
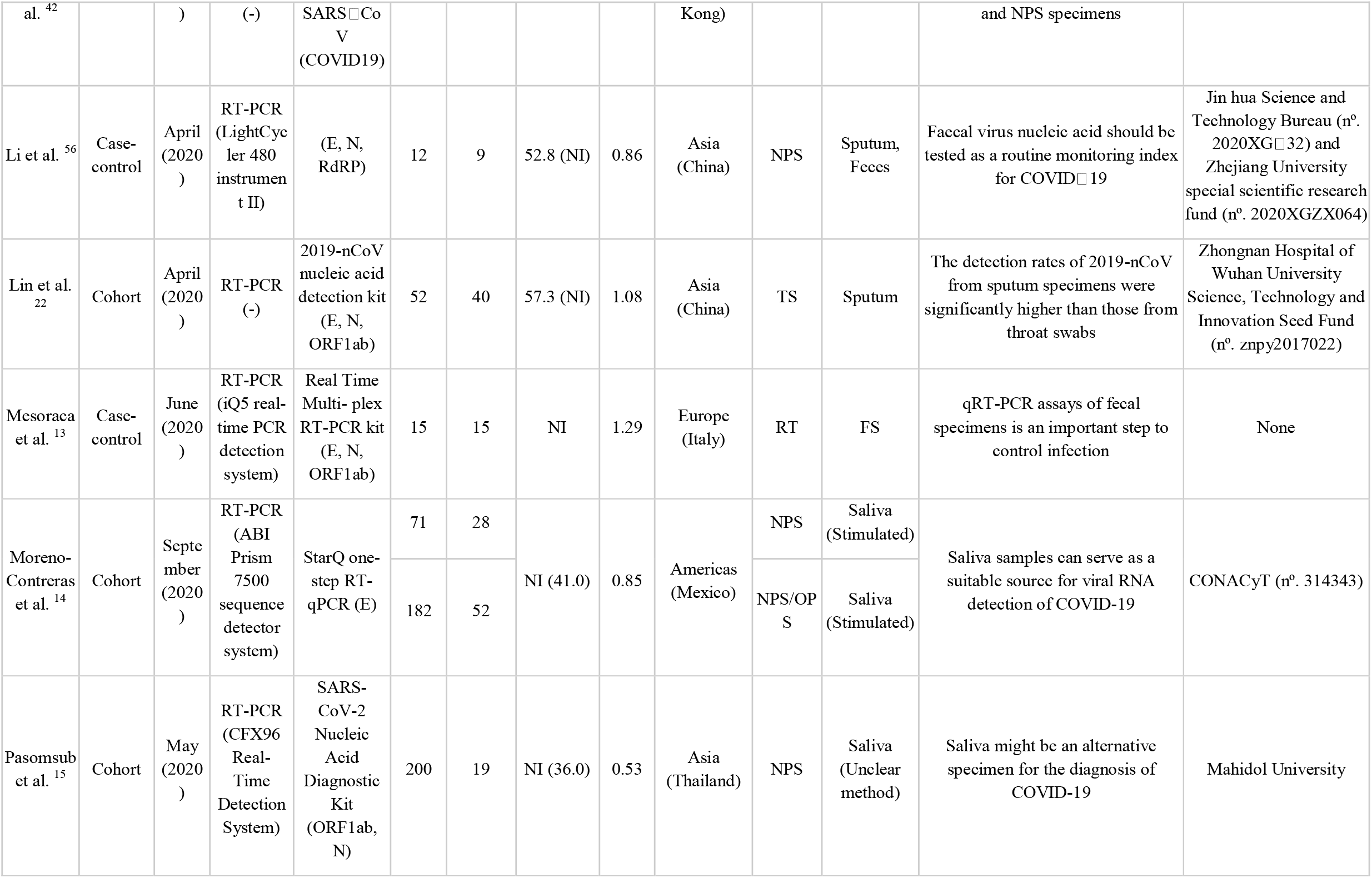
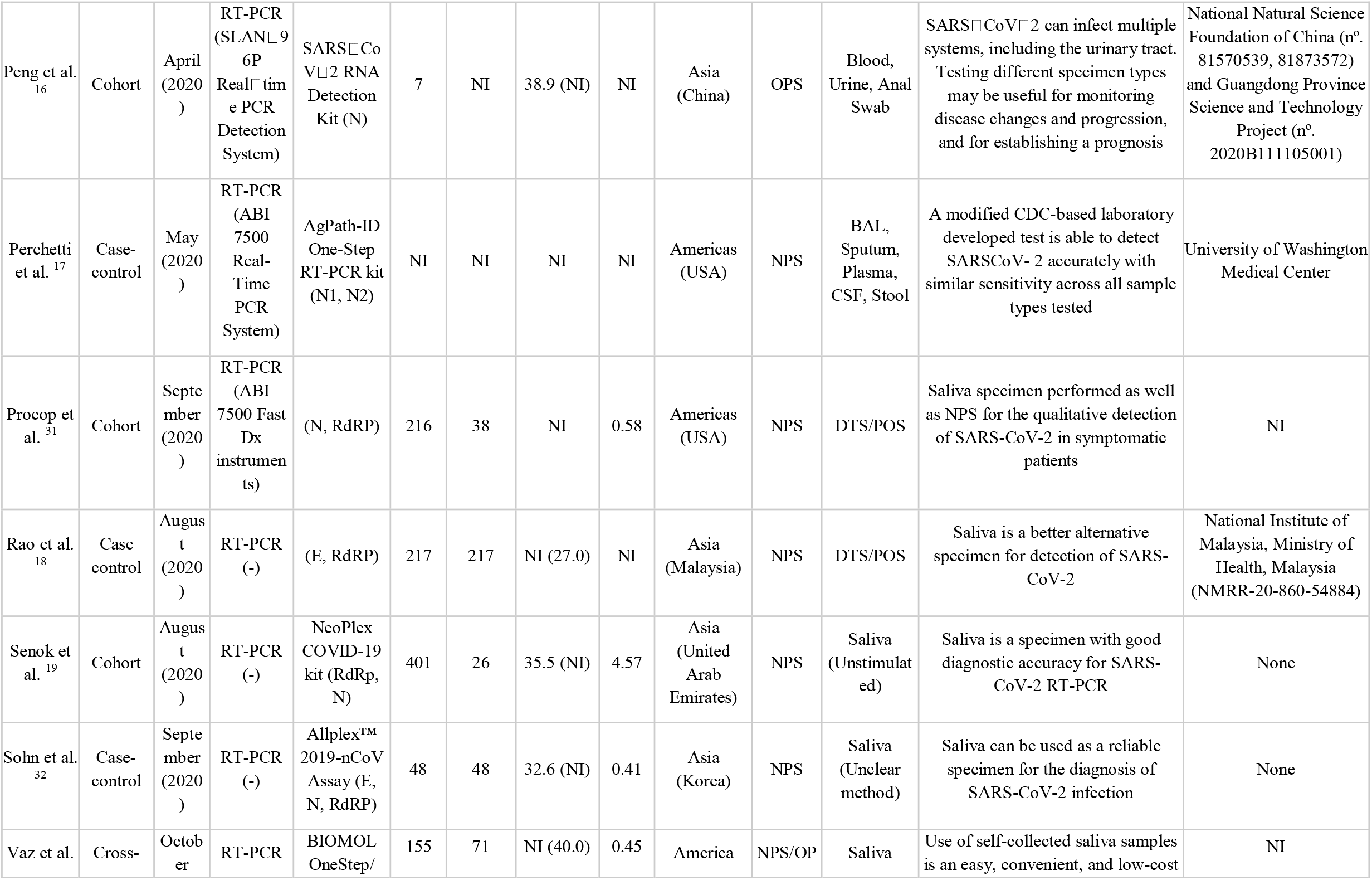
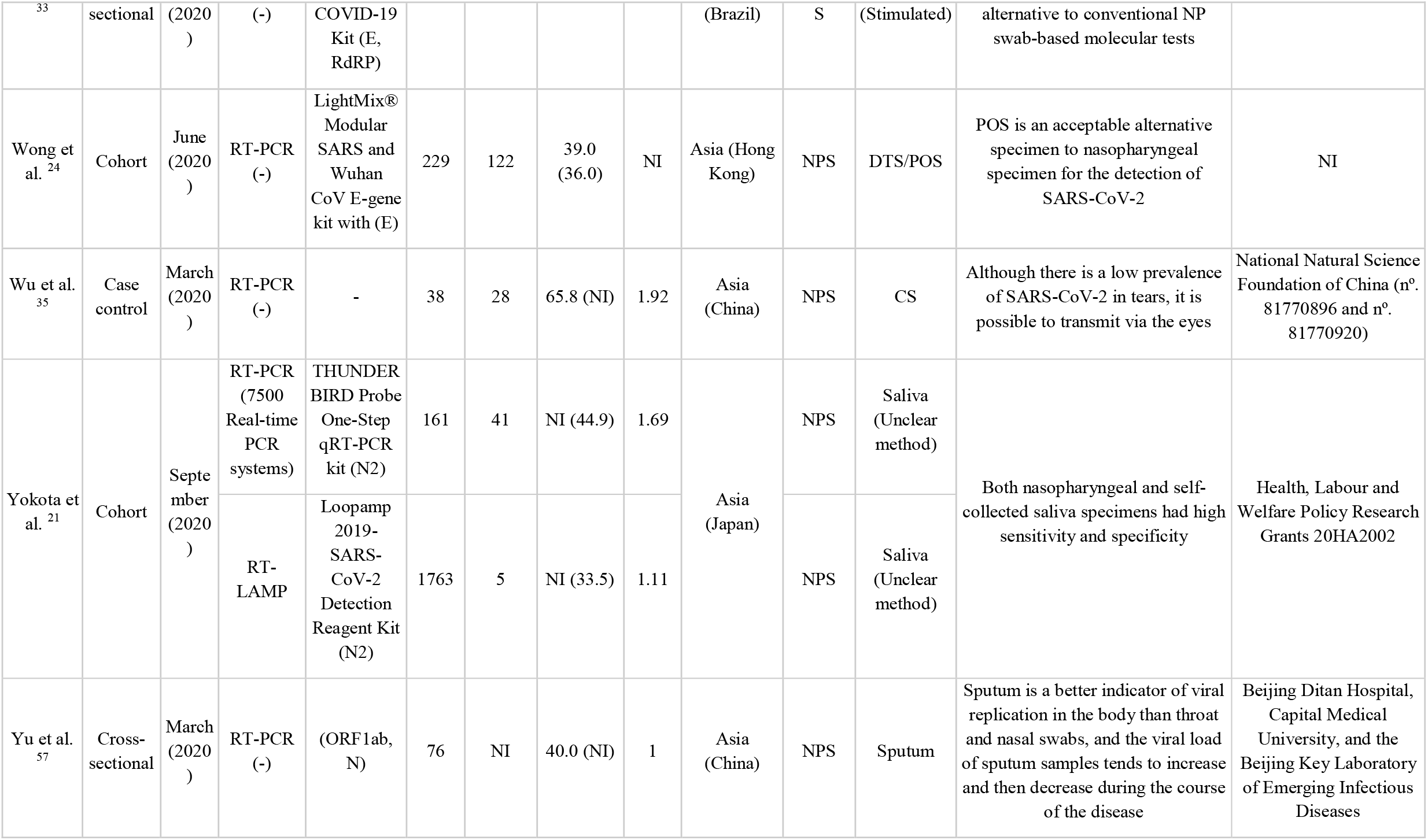
Characteristics of the included studies.

### 3.2. Quality Assessment

Overall, twenty-one studies had low risk of bias (63.6%) ^11–24,34,37–42^, eleven raised some concerns (33.3%) ^20,25–33,35^ and one had high risk of bias (3.0%) ^36^ (Figure 2) (fully detailed in appendix S3, pp 12). Some studies failed to provide information regarding index tests (33.3%, n=11), patient selection (12.1%, n=4) and reference standard (12.1%, n=4). Also, 36.4% (n=12), 15.2% (n=5) and 3.0% (n=1) of the studies raised some concerns regarding flow and timing, index test and reference standards, respectively. Out of the total (3.0%), one single study ^36^ was found to have a high risk of bias on “patient selection” and the “flow and timing” domains.

**Figure 2.**
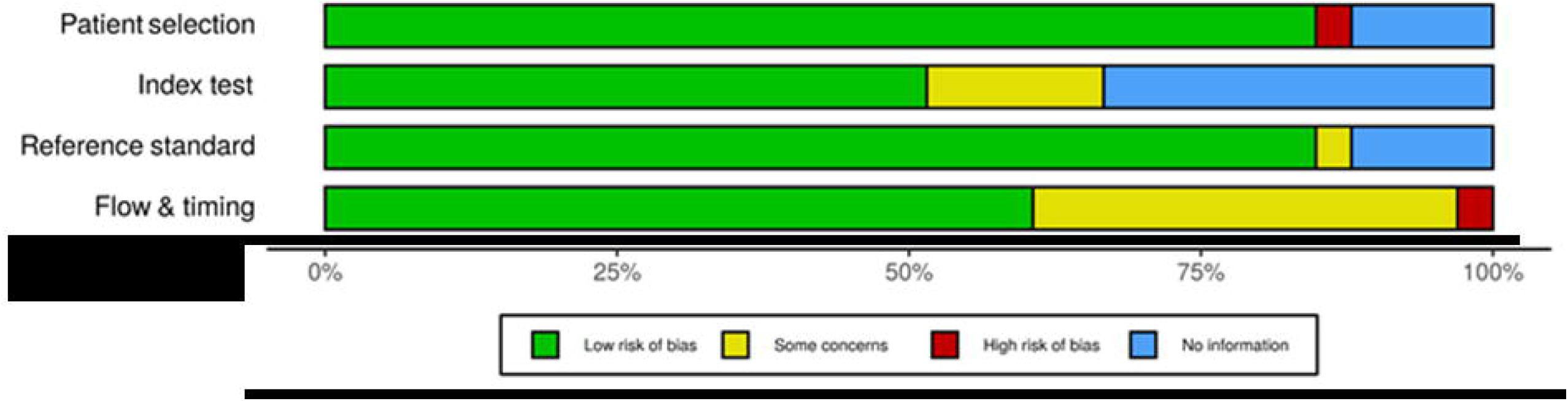
Summary of the risk of bias of the included studies (QUADAS-2).

### 3.3. Quantitative Analysis (Meta-analysis)

Random-effects meta-analysis demonstrated saliva as the index specimen with higher sensitivity and lower false-positive rate test results (Table 2).

In the meta-analysis of salivary samples from the oral cavity, estimates show an overall diagnostic accuracy of 92.1% (figure 3a; 0.921, 95% CI: 0.700,0.983), with an estimated sensitivity of 83.9% (figure 3b; 0.839, 95% CI: 0.774;0.888) and specificity of 96.4% (figure 3c; 0.964, 95% CI: 0.895;0.988).

**Table 2.**
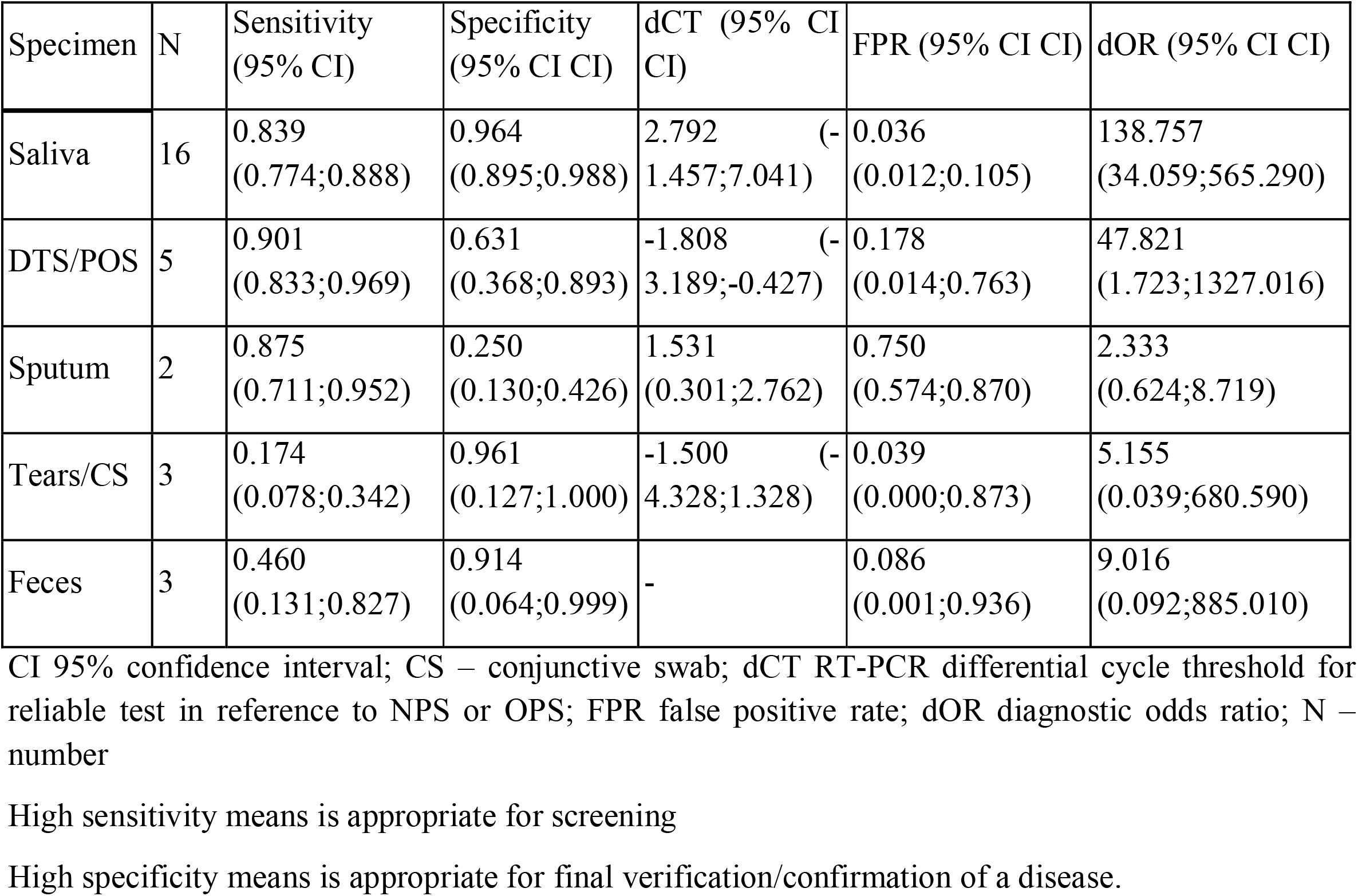
Estimated diagnostic parameters for different specimens.

**Figure 3.**
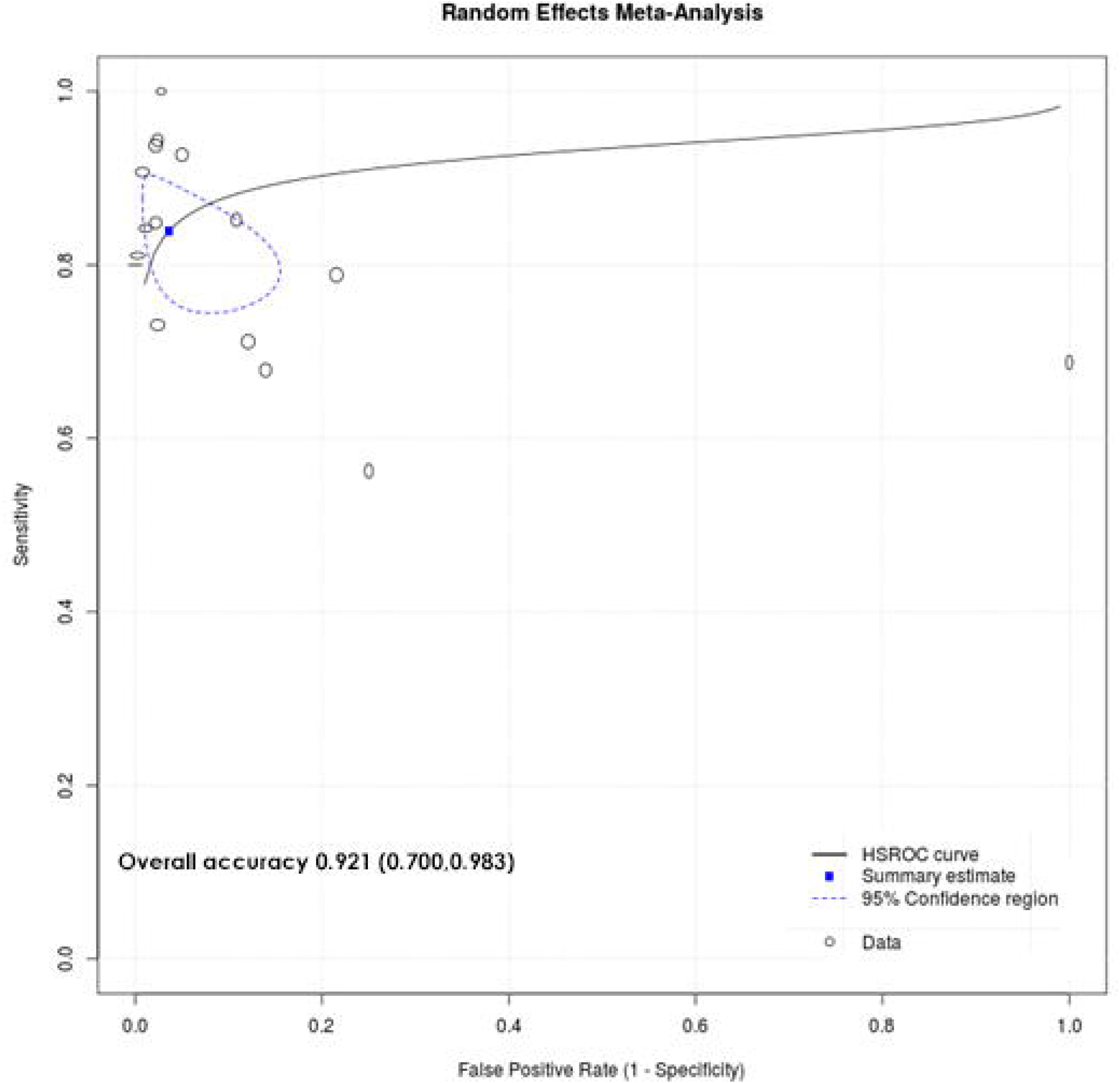
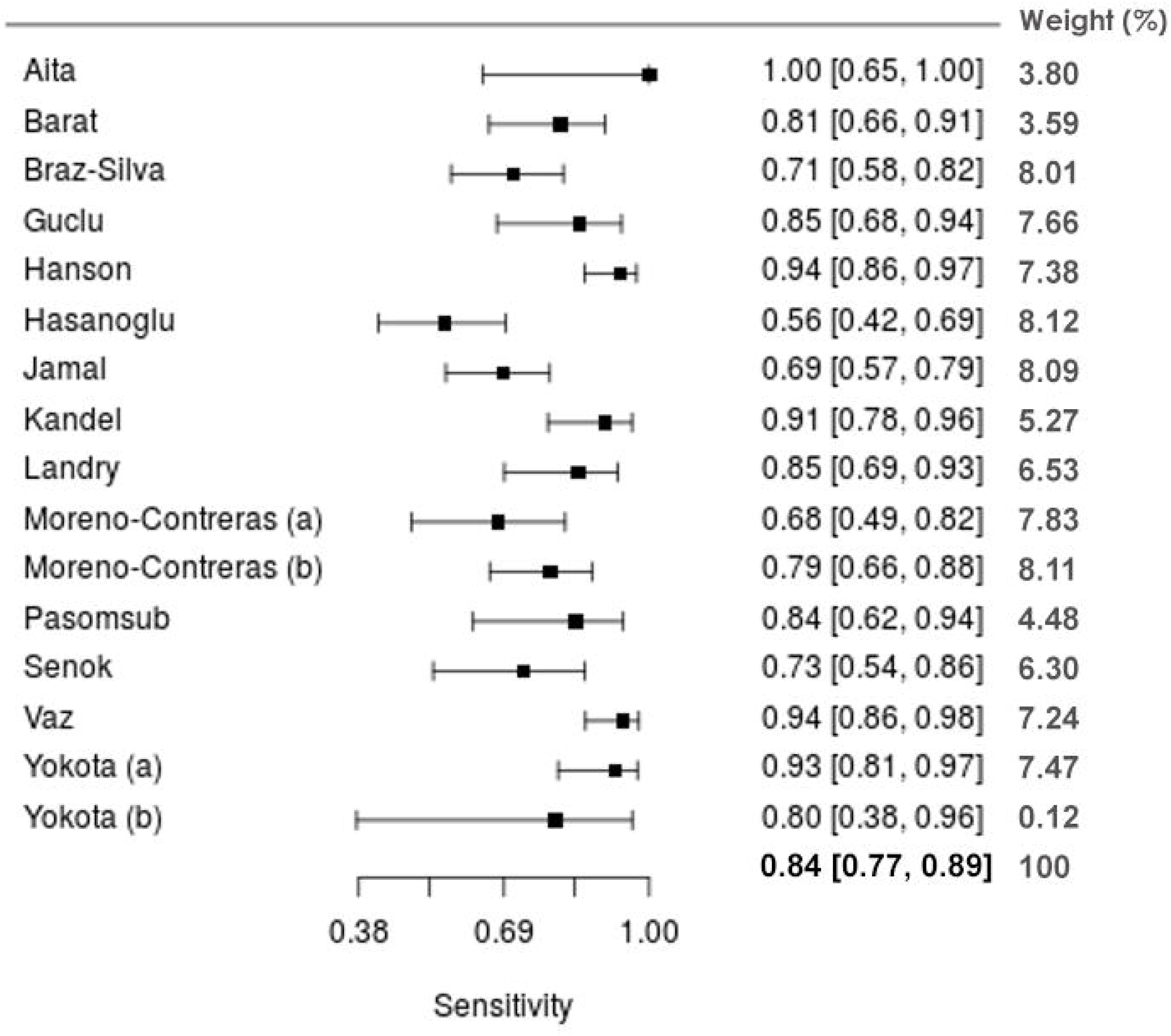
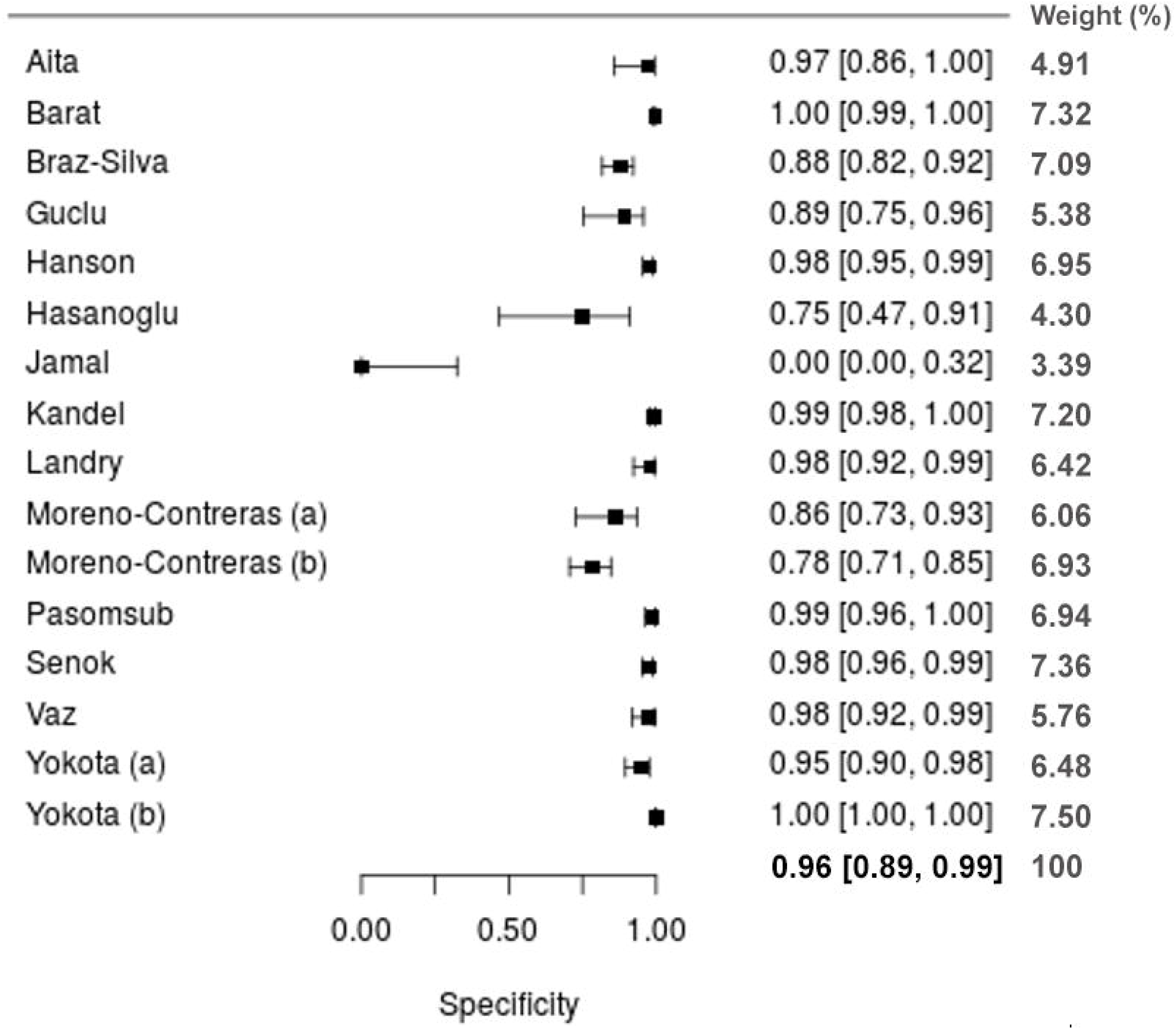
Meta-analytical estimates for saliva. (a) sROC plots with a curve (solid line), 95% confidence region (dashed line), summary point (blue square) (and every circle represents the sensitivity and specificity estimate from one study, and the size of the circle reflects the relative weight); (b) forest plot of the sensitivity; (c) forest plot of the specificity.

Meta-regressions screening for potential confounding variables demonstrate no influence of M/F ratio (appendix S4, pp 13). Regarding the differences in study sample size, while for sensitivity it is not significant (p=0.518) (Figure S7), for specificity a higher sample size appears to impact positively its performance (p<0.034) (appendix S4, pp 13). As for the target gene, sub-analysis was deemed unsuitable given the variety of methods (Table 1).

Concerning meta-analysis of DTS/POS based tests, estimates show an overall diagnostic accuracy of 79.7% (figure 4a; 0.797, 95% CI: 0.433;0.953), with an estimated sensitivity of 90.1% (figure 4b; 0.901, 95% CI: 0.833;0.969) and specificity of 63.1% (figure 4c; 0.631, 95% CI: 0.368;0.893). The uncertainty of test performance estimates is much higher than in saliva-based diagnostics since less studies support the meta-analysis model fit. Meta-regression suggests that M/F ratio have a negative confounding effect on test specificity (p<0.001) (appendix S4, pp 13). Estimates concerning sputum show an overall diagnostic sensitivity and specificity of 85.4% (0.875, 95% CI: 0.711,0.952) and 25.4% (0.250, 95% CI: 0.130,0.426), respectively. Due to the low number of studies available (n=2), the sROC analysis was not performed.

**Figure 4.**
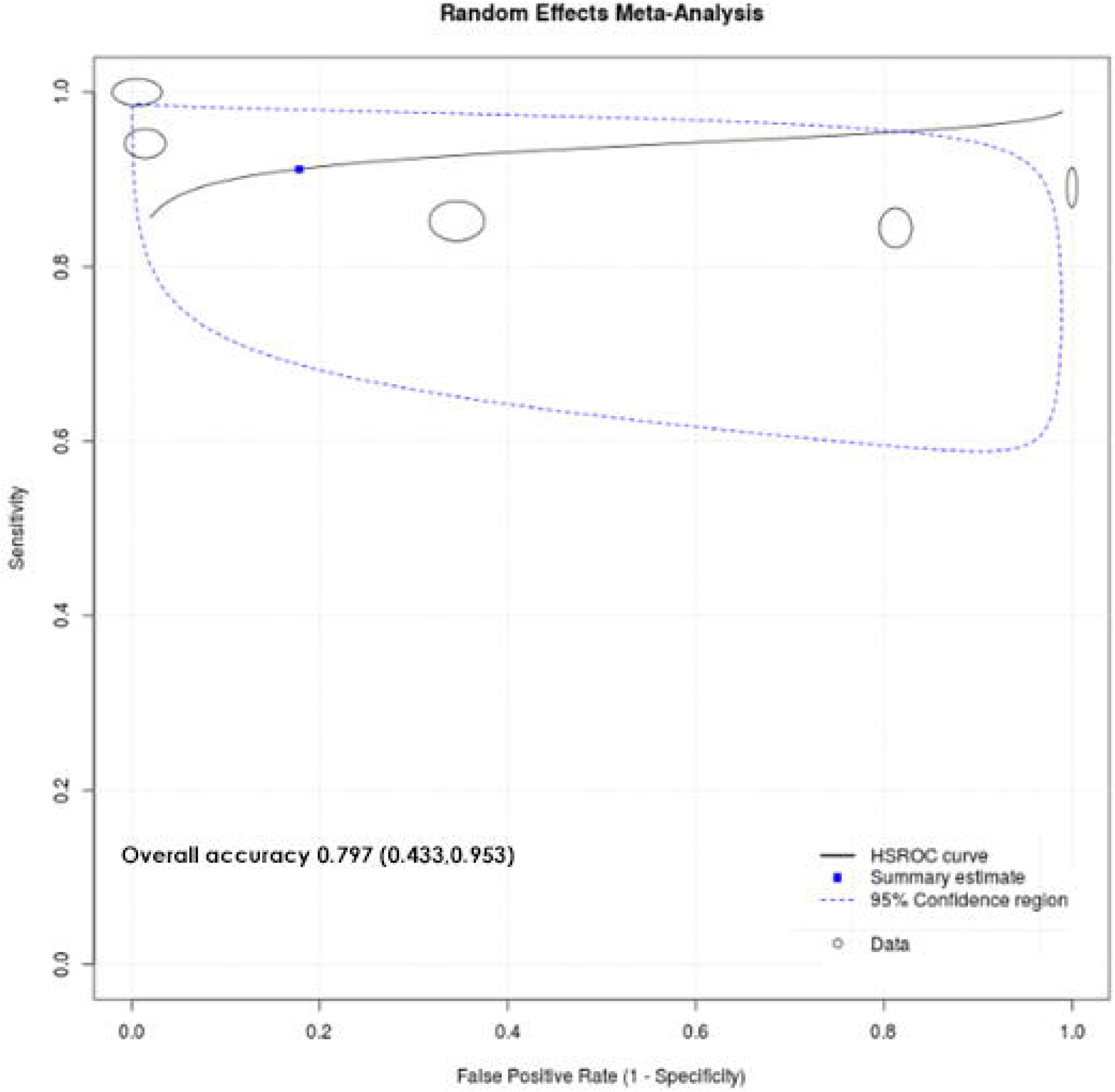
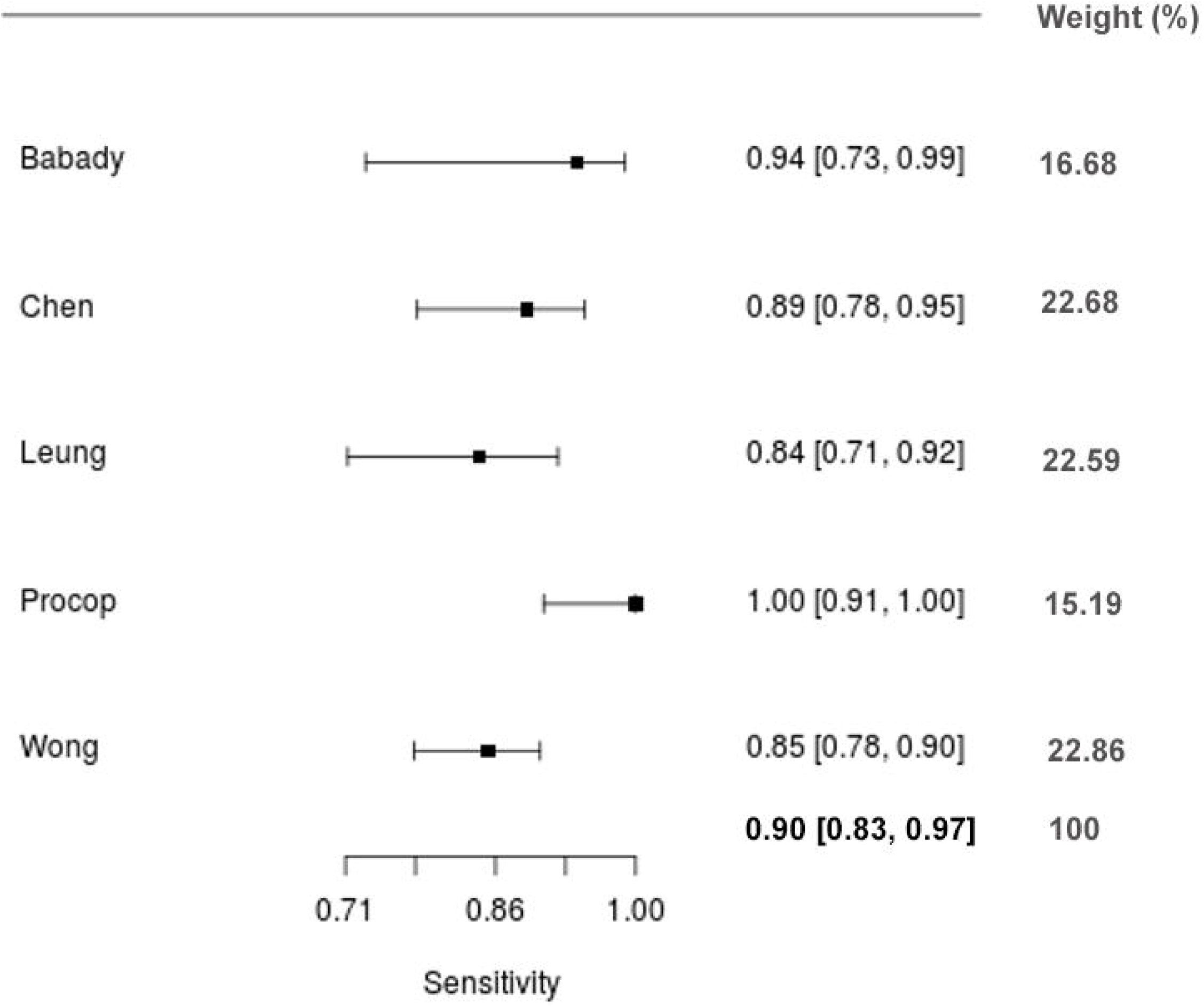
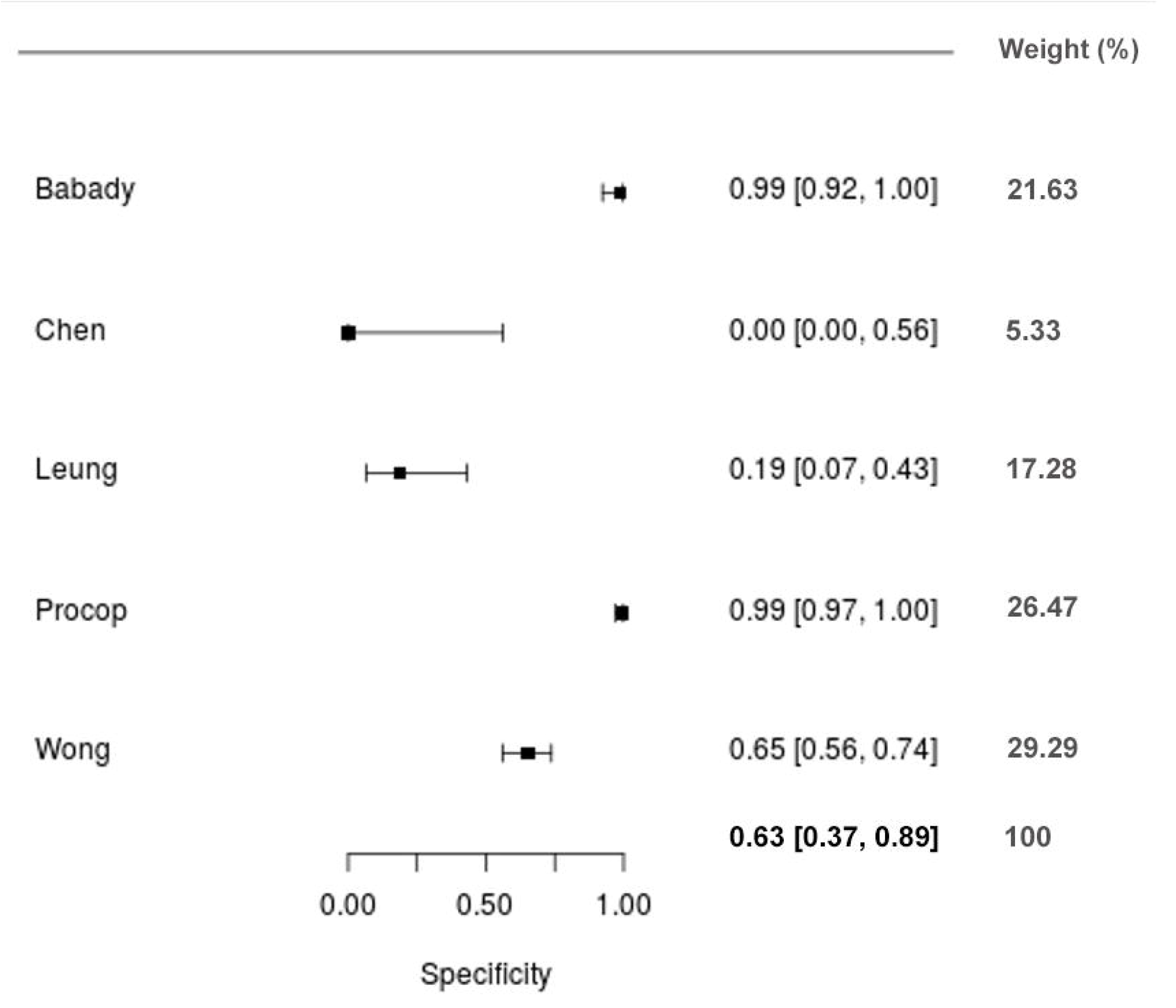
Meta-analytical estimates for DTS/POS. (a) sROC plots with a curve (solid line), 95% confidence region (dashed line), summary point (blue square) (and every circle represents the sensitivity and specificity estimate from one study, and the size of the circle reflects the relative weight); (b) forest plot of the sensitivity; (c) forest plot of the specificity.

Studies on tears/CS had an overall sensitivity of 17.4% (0.174, 95% CI: 0.078,0.342) and an overall specificity of 96.1% (0.961, 95% CI: 0.127,1.000) (Table 2). Meta-regressions showed that specificity has a positive correlation with M/F ratio (p=0.037) (appendix S4, pp 13).

In what concerns feces/anal swab, the overall diagnostic sensitivity was 46.0% (0.460, 95% CI: 0.131,0.827) while the overall specificity was 91,4% (0.914, 95% CI: 0.064,0.999) (Table 2) (appendix S7-S8, pp 13-14). Meta-regressions show no confounding variables towards the performance results (appendix S4, pp 13).

Regarding urine, we found not enough studies to compute estimates.

Finally, the cycle threshold (CT) in RT-PCR tests was compared between the index samples under analysis. We obtained an overall mean difference between saliva and NPS/OPS of 2.792 (95% CI: −1.457;7.041) (appendix S9, pp 14), i.e., there is a negative correlation between the CT for the NPS/OPS specimen and the CT for saliva samples. This means that, on average, the CT value for saliva is higher than the one for NPS/OPS. For the mean difference between DTS/POS and NPS/OPS, a significantly different estimate was obtained: −1.808 (95% CI: −3.189;-0.427) (appendix S10, pp 14).

## 4. Discussion

We systematically reviewed studies on the diagnostic accuracy of RT-PCR testing using minimally invasive human specimens that may replace nasal and throat swabbing, which are routinely used for the detection of SARS-CoV-2. Overall, the most promising index specimen is saliva, with a true positive rate (sensitivity-pooled estimate) of 83.9%) and a true negative rate (specificity-pooled estimate) above 90% (96.4%). Interestingly, a critical analysis of these results shows that the tests’ accuracy was affected by a high level of heterogeneity, mostly due to methodological variations. Therefore, as a diagnostic specimen, “saliva” deserves a particular attention, and several considerations will be made. Firstly, most studies accounted for salivary samples circumscribed to the oral region (anterior to the throat) ^11,12,14,19,21,25,29,32–34,36–39,41^, while the remaining studies analysed DTS/POS with or without pre-throat saliva ^24,26,28,31,40,42^. This fact is very important as the salivary characteristics and the collection method differ, and the DTS/POS may contain samples other than the oropharyngeal region (naso-pharyngeal or laryngeal-pharyngeal) ^52^. Secondly, among saliva samples from the oral cavity, the described methods show high heterogeneity and are unclear, as is the case regarding whether saliva was stimulated or not. Nevertheless, despite the multiple approaches used for salivary collection from the oral cavity (stimulated, unstimulated or unclear), saliva provided a high diagnostic accuracy (above 90%), confirming the potential of this specimen for SARS-Cov-2 detection. An additional limitation is that some of these works failed to properly describe the percentage of patients having asymptomatic, pre-symptomatic or symptomatic statuses, as the VL varies significantly in these patients and may negatively affect the accuracy of saliva as an index specimen. To further improve the saliva collecting protocol and secure its clinical validation and utility, specifically designed studies shall be performed, overcoming current methodological limitations.

Concerning the other evaluated index specimens, sputum presented an elevated risk of delivering false positive results when compared to NPS/OPS RT-PCR. Nevertheless, we must be cautious in interpreting these results due to the small number of studies. Similarly, tears/CS delivered the lowest sensitivity and yet, the highest specificity though, once again, these results were based on scarce data.

As for the CT analyses, due to the low number of available studies, these estimates are inconclusive at this stage.

From the sampling standpoint, both saliva and sputum can be easily collected; however, 72% of COVID-19 patients may not produce enough sputum for analysis ^53^. Therefore, saliva (oral region) seems to be the best specimen for both public health and epidemiologic measures ^52^. Because saliva can be self-collected by patients at home or the outbreak spot, it would decrease the exposure of health-care workers to infections, and reduce the waiting times for sample collection ^52^. On the contrary, DTS/POS may cause the dispersion of aerosols as a result of the cough up collection process. Nevertheless, some papers reported lower accuracy scores in salivary samples owing to critical factors such as the viral load ^54^, which greatly depend on the disease stage (time from onset of illness) and the time-point of specimen collection over the day. This is why, the influence of the specific time of sampling and the disease stage on the accuracy rate of the test were both considered in this systematic review through meta-regression, although unsuccessfully. Given this, future studies should focus on these factors in order to enable more concluding results and eventually define a detailed protocol for sampling prior to collection (e.g. timepoint, oral hygiene, avoid drinking or eating beforehand, physical exercise).

We are unaware of any other similar systematic review pooling consistent estimates on alternative specimens to detect SARS-CoV-2, in a way that it could have a significant impact in the accepted sampling methodologies. Indeed, almost ten months have passed since the public announcement of the COVID-19 pandemic and we have now access to a large number of scientific articles. The timing of this review is thus adequate and decisive to ensure the computation of pooling estimates, which, nonetheless, might turn outdated in the next months. Notwithstanding, these results pinpoint saliva sample circumscribed to the oral cavity as the index specimen with the greatest potential. This is a very important outcome owing to the particular circumstances we are currently experiencing (second or third waves of COVID-19) demanding the massive and rapid diagnosis of infection for which a self-administrated protocol for specimen collection would be extremely useful.

### Strengths and Limitations

Despite the thorough and comprehensive approach undertaken in this review to appraise all clinical evidence, some shortcomings deserve attention. The high level of heterogeneity observed limits the validation of quantitative analyses. This might be explained by the methodological variability among works, namely the diverse number of samples considered in each work, not all studies have followed the same index test, and the sample treatment and the target gene used was not always the same.

Many studies addressed the topic of detecting the presence of SARS-CoV-2 in index samples but not all of them could be included in this meta-analysis since some of them did not provide all the raw data required to calculate the main diagnostic performance parameters. Moreover, some of the works only tested positive patients. Other factors that can induce some variance in results are the timing of specimens’ collection and testing, sample treatment, among others, and some of the publications did not provide such information. Given the urgency to provide solutions for the COVID-19 pandemic, this heterogeneity might be seen as a collateral limitation.

These results resulted from a rigorous protocol with up-to-date standards using appropriate guidelines and we were also able to estimate the accuracy (clinical sensitivity and specificity) for a considerable number of index specimens. Despite the urgent need for better designed trials, with more homogeneous methodologies to further confirm our findings, in the short-run, they may aid public health authorities in deciding to use alternative samples for SARS-Cov-2 infection diagnosis which are as reliable as nasal and throat swabs, but are non-painful, non-stressful and much easier to collect.

## 5. Conclusions

Despite having several vaccines against SARS-CoV-2 already approved and being implemented in most developed countries, the coverage has been very slow, and it will take months to significantly reduce the prevalence of COVID-19. Since the very beginning of the pandemic, massive testing has been a critical priority in the struggle against the spread of the virus. Effective tests allow to discriminate between infected and non-infected people, thereby supporting decision making for clinical management of patients, transmission control, and epidemiological studies. According to the WHO interim guidance regarding “Laboratory testing guiding principles” ^55^, the availability of accurate laboratory or point-of-care tests are as important as the rapid collection of appropriate physiological samples. Respiratory specimens are the only ones that were accepted up to now, but the complexity in their collection from the nasal cavity and discomfort caused to patients are driving the search for simpler and less intrusive alternatives.

To this end, several alternative specimens have been compared to nasal/throat swabs for diagnosis of SARS-Cov-2 infection using nucleic acid assays (RT-PCR), and the results were systematically reviewed herein. We found that saliva from the oral region is the best candidate as an alternative specimen for SARS-Cov-2 detection. In fact, despite some heterogeneity in methodologies, the proportion of infected and non-infected patients correctly identified through the index sample is 83.9%, and 96.4%, respectively. The second-best specimen was DTS/POS, with a better true positive rate than saliva (sensitivity of 90.1%), but a much lower true negative rate (specificity of 63.1%). The specificity of sputum samples was even lower (25.4%), despite a reasonably high sensitivity (85.4%). Globally, the clinical performance of the other specimens (urine, feces, and tears) was inferior, but one should mention that the number of studies with these index specimens done so far is still scarce.

To sum up, saliva samples simply taken from the oral cavity are promising alternatives to the currently used swab-based specimens, since they can be effective, and allow self-collection. Besides mitigating the discomfort caused by sampling, saliva testing may considerably reduce the transmission risk while increasing testing capacity, ultimately promoting the implementation of truly deployable COVID-19 tests, which could either work at the point-of-care (e.g. hospitals, clinics) or outbreak control spots (e.g. schools, airports, and nursing homes). Before the index specimen saliva can be recommended by the main public health authorities, further assessment and validation is urgently required to define the best practices to adopt.

## Data Availability

Data will be provided upon reasonable request

## Contributors

VMM, PM, VM, JB and MGA designed the study and wrote the draft manuscript. VMM and PM did the systematic reviews. VMM and PM searched the literature. VM, JB, JJM, NT and MGA actively discussed and provided insightful suggestions. All authors critically reviewed the methods and results and contributed to writing the article.

## Data sharing

All data is presented in the manuscript and in the supplementary document available online.

## Acknowledgments and funding

This work was supported by national funds from FCT -Foundation for Science and Technology, I.P. through the CiiEM (project IDB/04585/2020), the Applied Molecular Biosciences Unit-UCIBIO (project UID/Multi/04378/2013), co-financed by the ERDF under the PT2020 Partnership Agreement (POCI-01-0145-FEDER-007728) and the program Research4COVID 19 (Project nr. 662).

