## appendix for "Diagnosis of SARS-Cov-2 infection using specimens other than naso- and oropharyngeal swabs: a systematic review and meta-analysis"

***Systematic Review***

**Supplementary Files**

**Title Physiological specimens collected with no use of nasal and throat swabs for COVID-19 testing: a systematic review and meta-analysis**

Vânia M. Moreira ^1^, Paulo Mascarenhas ^2,3^, Vanessa Machado ^2,3^, João Botelho ^2,3^, José João Mendes ^2,3^ and M. Gabriela Almeida ^2,4,^*

^1^ Área departamental de Engenharia Química, Instituto Superior de Engenharia de Lisboa; Rua Conselheiro Emídio Navarro, 1 1959-007 Lisboa, Portugal

^2^	Centro de Investigação Interdisciplinar Egas Moniz (CiiEM), Egas Moniz - Cooperativa de Ensino Superior CRL, Campus Universitário, Quinta da Granja, 2829-511 Caparica, Portugal

^3^	Evidence-Based Hub, CiiEM, Egas Moniz - Cooperativa de Ensino Superior CRL, Campus Universitário, Quinta da Granja, 2829-511 Caparica, Portugal

^4^	UCIBIO, REQUIMTE, Faculdade de Ciências e Tecnologia, Universidade Nova de Lisboa, 2829-516 Monte de Caparica, Portugal

### **Appendix S1**. PRISMA 2009 Checklist

| **Section/topic** | **#** | **Checklist item** | **Reported on page #** |
| --- | --- | --- | --- |
| **TITLE** | | |  |
| Title | 1 | Identify the report as a systematic review, meta-analysis, or both. |  |
| **ABSTRACT** | | |  |
| Structured summary | 2 | Provide a structured summary including, as applicable: background; objectives; data sources; study eligibility criteria, participants, and interventions; study appraisal and synthesis methods; results; limitations; conclusions and implications of key findings; systematic review registration number. |  |
| **INTRODUCTION** | | |  |
| Rationale | 3 | Describe the rationale for the review in the context of what is already known. |  |
| Objectives | 4 | Provide an explicit statement of questions being addressed with reference to participants, interventions, comparisons, outcomes, and study design (PICOS). |  |
| **METHODS** | | |  |
| Protocol and registration | 5 | Indicate if a review protocol exists, if and where it can be accessed (e.g., Web address), and, if available, provide registration information including registration number. |  |
| Eligibility criteria | 6 | Specify study characteristics (e.g., PICOS, length of follow-up) and report characteristics (e.g., years considered, language, publication status) used as criteria for eligibility, giving rationale. |  |
| Information sources | 7 | Describe all information sources (e.g., databases with dates of coverage, contact with study authors to identify additional studies) in the search and date last searched. |  |
| Search | 8 | Present full electronic search strategy for at least one database, including any limits used, such that it could be repeated. |  |
| Study selection | 9 | State the process for selecting studies (i.e., screening, eligibility, included in systematic review, and, if applicable, included in the meta-analysis). |  |
| Data collection process | 10 | Describe method of data extraction from reports (e.g., piloted forms, independently, in duplicate) and any processes for obtaining and confirming data from investigators. |  |
| Data items | 11 | List and define all variables for which data were sought (e.g., PICOS, funding sources) and any assumptions and simplifications made. |  |
| Risk of bias in individual studies | 12 | Describe methods used for assessing risk of bias of individual studies (including specification of whether this was done at the study or outcome level), and how this information is to be used in any data synthesis. |  |
| Summary measures | 13 | State the principal summary measures (e.g., risk ratio, difference in means). |  |
| Synthesis of results | 14 | Describe the methods of handling data and combining results of studies, if done, including measures of consistency (e.g., I^2^) for each meta-analysis. |  |

| **Section/topic** | **#** | **Checklist item** | **Reported on page #** |
| --- | --- | --- | --- |
| Risk of bias across studies | 15 | Specify any assessment of risk of bias that may affect the cumulative evidence (e.g., publication bias, selective reporting within studies). |  |
| Additional analyses | 16 | Describe methods of additional analyses (e.g., sensitivity or subgroup analyses, meta-regression), if done, indicating which were pre-specified. |  |
| **RESULTS** | | |  |
| Study selection | 17 | Give numbers of studies screened, assessed for eligibility, and included in the review, with reasons for exclusions at each stage, ideally with a flow diagram. |  |
| Study characteristics | 18 | For each study, present characteristics for which data were extracted (e.g., study size, PICOS, follow-up period) and provide the citations. |  |
| Risk of bias within studies | 19 | Present data on risk of bias of each study and, if available, any outcome level assessment (see item 12). |  |
| Results of individual studies | 20 | For all outcomes considered (benefits or harms), present, for each study: (a) simple summary data for each intervention group (b) effect estimates and confidence intervals, ideally with a forest plot. |  |
| Synthesis of results | 21 | Present results of each meta-analysis done, including confidence intervals and measures of consistency. |  |
| Risk of bias across studies | 22 | Present results of any assessment of risk of bias across studies (see Item 15). |  |
| Additional analysis | 23 | Give results of additional analyses, if done (e.g., sensitivity or subgroup analyses, meta-regression [see Item 16]). |  |
| **DISCUSSION** | | |  |
| Summary of evidence | 24 | Summarize the main findings including the strength of evidence for each main outcome; consider their relevance to key groups (e.g., healthcare providers, users, and policy makers). |  |
| Limitations | 25 | Discuss limitations at study and outcome level (e.g., risk of bias), and at review-level (e.g., incomplete retrieval of identified research, reporting bias). |  |
| Conclusions | 26 | Provide a general interpretation of the results in the context of other evidence, and implications for future research. |  |
| **FUNDING** | | |  |
| Funding | 27 | Describe sources of funding for the systematic review and other support (e.g., supply of data); role of funders for the systematic review. |  |

NA – Not applicable

*From:* Moher D, Liberati A, Tetzlaff J, Altman DG, The PRISMA Group (2009). Preferred Reporting Items for Systematic Reviews and Meta-Analyses: The PRISMA Statement. PLoS Med 6(7): e1000097. doi:10.1371/journal.pmed1000097

For more information, visit: **www.prisma-statement.org**.

### **Appendix S2**. List of potentially relevant studies not included in the systematic review, along with the reasons for exclusion.

| **#** | **Reference** | **Reason for exclusion** |
| --- | --- | --- |
| 1 | Alsuliman T, Sulaiman R, Ismail S, Srour M, Alrstom A. COVID-19 paraclinical diagnostic tools: Updates and future trends. Curr Res Transl Med. 2020 Aug;68(3):83-91. doi: 10.1016/j.retram.2020.06.001. | Review |
| 2 | Altawalah H, AlHuraish F, Alkandari WA, Ezzikouri S. Saliva specimens for detection of severe acute respiratory syndrome coronavirus 2 in Kuwait: A cross-sectional study. J Clin Virol. 2020 Nov;132:104652. doi: 10.1016/j.jcv.2020.104652. | Unclear results |
| 3 | Arora R, Goel R, Kumar S, Chhabra M, Saxena S, Manchanda V, Pumma P. Evaluation of SARS-CoV-2 in Tears of Patients with Moderate to Severe COVID-19. Ophthalmology. 2020 Aug 31:S0161-6420(20)30847-2. doi: 10.1016/j.ophtha.2020.08.029. | No control |
| 4 | Azzi L, Carcano G, Gianfagna F, Grossi P, Gasperina DD, Genoni A, Fasano M, Sessa F, Tettamanti L, Carinci F, Maurino V, Rossi A, Tagliabue A, Baj A. Saliva is a reliable tool to detect SARS-CoV-2. J Infect. 2020 Jul;81(1):e45-e50. doi: 10.1016/j.jinf.2020.04.005. | Review |
| 5 | Bullis SSM, Crothers JW, Wayne S, Hale AJ. A cautionary tale of false-negative nasopharyngeal COVID-19 testing. IDCases. 2020 May 5;20:e00791. doi: 10.1016/j.idcr.2020.e00791. | Case report |
| 6 | Bwire GM, Majigo MV, Njiro BJ, Mawazo A. Detection profile of SARS-CoV-2 using RT-PCR in different types of clinical specimens: A systematic review and meta-analysis. J Med Virol. 2020 Jul 24:10.1002/jmv.26349. doi: 10.1002/jmv.26349. | Review |
| 7 | Byrne RL, Kay GA, Kontogianni K, Aljayyoussi G, Brown L, Collins AM, Cuevas LE, Ferreira DM, Fraser AJ, Garrod G, Hill H, Hughes GL, Menzies S, Mitsi E, Owen SI, Patterson EI, Williams CT, Hyder-Wright A, Adams ER, Cubas-Atienzar AI. Saliva Alternative to Upper Respiratory Swabs for SARS-CoV-2 Diagnosis. Emerg Infect Dis. 2020 Nov;26(11):2770-2771. doi: 10.3201/eid2611.203283. | Letter to the editor |
| 8 | Cevik M, Bamford CGG, Ho A. COVID-19 pandemic-a focused review for clinicians. Clin Microbiol Infect. 2020 Jul;26(7):842-847. doi: 10.1016/j.cmi.2020.04.023. | Review |
| 9 | Chen L, Lou J, Bai Y, Wang M. COVID-19 Disease With Positive Fecal and Negative Pharyngeal and Sputum Viral Tests. Am J Gastroenterol. 2020 May;115(5):790. doi: 10.14309/ajg.0000000000000610. | Letter to the editor |
| 10 | Chen Y, Chen L, Deng Q, Zhang G, Wu K, Ni L, Yang Y, Liu B, Wang W, Wei C, Yang J, Ye G, Cheng Z. The presence of SARS-CoV-2 RNA in the feces of COVID-19 patients. J Med Virol. 2020 Jul;92(7):833-840. doi: 10.1002/jmv.25825. | No information regardinbg TP, FN, TN and FP |
| 11 | Chen Z, Tong L, Zhou Y, Hua C, Wang W, Fu J, Shu Q, Hong L, Xu H, Xu Z, Chen Y, Mao Y, Ye S, Wu X, Wang L, Luo Y, Zou X, Tao X, Zhang Y. Childhood COVID-19: a multicentre retrospective study. Clin Microbiol Infect. 2020 Sep;26(9):1260.e1-1260.e4. doi: 10.1016/j.cmi.2020.06.015. | One single samples |
| 12 | Cheng VCC, Wong SC, Chen JHK, Yip CCY, Chuang VWM, Tsang OTY, Sridhar S, Chan JFW, Ho PL, Yuen KY. Escalating infection control response to the rapidly evolving epidemiology of the coronavirus disease 2019 (COVID-19) due to SARS-CoV-2 in Hong Kong. Infect Control Hosp Epidemiol. 2020 May;41(5):493-498. doi: 10.1017/ice.2020.58. | Case report |
| 13 | Cho SM, Ha GY. A Case of COVID-19 in a 45-Day-Old Infant with Persistent Fecal Virus Shedding for More Than 12 Weeks. Yonsei Med J. 2020 Oct;61(10):901-903. doi: 10.3349/ymj.2020.61.10.901. | Case report |
| 14 | Chong CY, Kam KQ, Li J, Maiwald M, Loo LH, Nadua KD, Tan NWH, Yung CF, Thoon KC. Saliva is not a useful diagnostic specimen in children with Coronavirus Disease 2019. Clin Infect Dis. 2020 Sep 14:ciaa1376. doi: 10.1093/cid/ciaa1376. | Letter to the editor |
| 15 | COVID-19 Investigation Team. Clinical and virologic characteristics of the first 12 patients with coronavirus disease 2019 (COVID-19) in the United States. Nat Med. 2020 Jun;26(6):861-868. doi: 10.1038/s41591-020-0877-5. | Letter to the editor |
| 16 | Czumbel LM, Kiss S, Farkas N, Mandel I, Hegyi A, Nagy Á, Lohinai Z, Szakács Z, Hegyi P, Steward MC, Varga G. Saliva as a Candidate for COVID-19 Diagnostic Testing: A Meta-Analysis. Front Med (Lausanne). 2020 Aug 4;7:465. doi: 10.3389/fmed.2020.00465. | Review |
| 17 | Jones DL, Baluja MQ, Graham DW, Corbishley A, McDonald JE, Malham SK, Hillary LS, Connor TR, Gaze WH, Moura IB, Wilcox MH, Farkas K. Shedding of SARS-CoV-2 in feces and urine and its potential role in person-to-person transmission and the environment-based spread of COVID-19. Sci Total Environ. 2020 Dec 20;749:141364. doi: 10.1016/j.scitotenv.2020.141364. | Review |
| 18 | Sakanashi D, Asai N, Nakamura A, Miyazaki N, Kawamoto Y, Ohno T, Yamada A, Koita I, Suematsu H, Hagihara M, Shiota A, Kurumiya A, Sakata M, Kato S, Muramatsu Y, Koizumi Y, Kishino T, Ohashi W, Yamagishi Y, Mikamo H. Comparative evaluation of nasopharyngeal swab and saliva specimens for the molecular detection of SARS-CoV-2 RNA in Japanese patients with COVID-19. J Infect Chemother. 2021 Jan;27(1):126-129. doi: 10.1016/j.jiac.2020.09.027. | Only saliva samples |
| 19 | Dang Y, Liu N, Tan C, Feng Y, Yuan X, Fan D, Peng Y, Jin R, Guo Y, Lou J. Comparison of qualitative and quantitative analyses of COVID-19 clinical samples. Clin Chim Acta. 2020 Nov;510:613-616. doi: 10.1016/j.cca.2020.08.033. | No information regardinbg TP, FN, TN and FP |
| 20 | Du W, Yu J, Liu X, Chen H, Lin L, Li Q. Persistence of SARS-CoV-2 virus RNA in feces: A case series of children. J Infect Public Health. 2020 Jul;13(7):926-931. doi: 10.1016/j.jiph.2020.05.025. | Case report |
| 21 | Fakheran O, Dehghannejad M, Khademi A. Saliva as a diagnostic specimen for detection of SARS-CoV-2 in suspected patients: a scoping review. Infect Dis Poverty. 2020 Jul 22;9(1):100. doi: 10.1186/s40249-020-00728-w. | Review |
| 22 | Fan H, Yu X, Fu X, Zhu H, Lv Z, Yi W, Zhang Q. Clinical implications of different specimen types for nucleic acid testing in two cases of COVID-19. J Int Med Res. 2020 Aug;48(8):300060520949067. doi: 10.1177/0300060520949067. | Case report |
| 23 | Fukumoto T, Iwasaki S, Fujisawa S, Hayasaka K, Sato K, Oguri S, Taki K, Nakakubo S, Kamada K, Yamashita Y, Konno S, Nishida M, Sugita J, Teshima T. Efficacy of a novel SARS-CoV-2 detection kit without RNA extraction and purification. Int J Infect Dis. 2020 Sep;98:16-17. doi: 10.1016/j.ijid.2020.06.074. | Comparison between different methods |
| 24 | Gupta S, Parker J, Smits S, Underwood J, Dolwani S. Persistent viral shedding of SARS-CoV-2 in faeces - a rapid review. Colorectal Dis. 2020 Jun;22(6):611-620. doi: 10.1111/codi.15138. | Review |
| 25 | Kaya H, Çalışkan A, Okul M, Sarı T, Akbudak İH. Detection of SARS-CoV-2 in the tears and conjunctival secretions of Coronavirus disease 2019 patients. J Infect Dev Ctries. 2020 Sep 30;14(9):977-981. doi: 10.3855/jidc.13224. | Case series |
| 26 | Hamid H, Khurshid Z, Adanir N, Zafar MS, Zohaib S. COVID-19 Pandemic and Role of Human Saliva as a Testing Biofluid in Point-of-Care Technology. Eur J Dent. 2020 Jun 3. doi: 10.1055/s-0040-1713020. | Review |
| 27 | Han MS, Choi EH, Chang SH, Jin BL, Lee EJ, Kim BN, Kim MK, Doo K, Seo JH, Kim YJ, Kim YJ, Park JY, Suh SB, Lee H, Cho EY, Kim DH, Kim JM, Kim HY, Park SE, Lee JK, Jo DS, Cho SM, Choi JH, Jo KJ, Choe YJ, Kim KH, Kim JH. Clinical Characteristics and Viral RNA Detection in Children With Coronavirus Disease 2019 in the R | Comparison between symptomatic and asymptomatic patients |
| 28 | Han MS, Seong MW, Heo EY, Park JH, Kim N, Shin S, Cho SI, Park SS, Choi EH. Sequential Analysis of Viral Load in a Neonate and Her Mother Infected With Severe Acute Respiratory Syndrome Coronavirus 2. Clin Infect Dis. 2020 Nov 19;71(16):2236-2239. doi: 10.1093/cid/ciaa447. | Case series |
| 29 | Han MS, Seong MW, Kim N, Shin S, Cho SI, Park H, Kim TS, Park SS, Choi EH. Viral RNA Load in Mildly Symptomatic and Asymptomatic Children with COVID-19, Seoul, South Korea. Emerg Infect Dis. 2020 Oct;26(10):2497-2499. doi: 10.3201/eid2610.202449. | Letter to the editor |
| 30 | Harikrishnan P. Saliva as a Potential Diagnostic Specimen for COVID-19 Testing. J Craniofac Surg. 2020 Sep;31(6):e653-e655. doi: 10.1097/SCS.0000000000006724. | No results |
| 31 | Hinojosa-Velasco A, de Oca PVB, García-Sosa LE, Mendoza-Durán JG, Pérez-Méndez MJ, Dávila-González E, Ramírez-Hernández DG, García-Mena J, Zárate-Segura P, Reyes-Ruiz JM, Bastida-González F. A case report of newborn infant with severe COVID-19 in Mexico: Detection of SARS-CoV-2 in human breast milk and stool. Int J Infect Dis. 2020 Nov;100:21-24. doi: 10.1016/j.ijid.2020.08.055. | Case report |
| 32 | Hu X, Deng Q, Li J, Chen J, Wang Z, Zhang X, Fang Z, Li H, Zhao Y, Yu P, Li W, Wang X, Li S, Zhang L, Hou T. Development and Clinical Application of a Rapid and Sensitive Loop-Mediated Isothermal Amplification Test for SARS-CoV-2 Infection. mSphere. 2020 Aug 26;5(4):e00808-20. doi: 10.1128/mSphere.00808-20. | Comparison between different methods |
| 33 | Huang CG, Lee KM, Hsiao MJ, Yang SL, Huang PN, Gong YN, Hsieh TH, Huang PW, Lin YJ, Liu YC, Tsao KC, Shih SR. Culture-Based Virus Isolation To Evaluate Potential Infectivity of Clinical Specimens Tested for COVID-19. J Clin Microbiol. 2020 Jul 23;58(8):e01068-20. doi: 10.1128/JCM.01068-20. | Genetic comparation |
| 34 | Huang JT, Ran RX, Lv ZH, Feng LN, Ran CY, Tong YQ, Li D, Su HW, Zhu CL, Qiu SL, Yang J, Xiao MY, Liu MJ, Yang YT, Liu SM, Li Y. Chronological Changes of Viral Shedding in Adult Inpatients With COVID-19 in Wuhan, China. Clin Infect Dis. 2020 Nov 19;71(16):2158-2166. doi: 10.1093/cid/ciaa631. | Fecal and urine samples mixed in the same analysis |
| 35 | Inomata T, Kitazawa K, Kuno T, Sung J, Nakamura M, Iwagami M, Takagi H, Midorikawa-Inomata A, Zhu J, Fujimoto K, Okumura Y, Miura M, Fujio K, Hirosawa K, Akasaki Y, Kuwahara M, Dana R, Murakami A. Clinical and Prodromal Ocular Symptoms in Coronavirus Disease: A Systematic Review and Meta-Analysis. Invest Ophthalmol Vis Sci. 2020 Aug 3;61(10):29. doi: 10.1167/iovs.61.10.29. | Review |
| 36 | Iwasaki S, Fujisawa S, Nakakubo S, Kamada K, Yamashita Y, Fukumoto T, Sato K, Oguri S, Taki K, Senjo H, Sugita J, Hayasaka K, Konno S, Nishida M, Teshima T. Comparison of SARS-CoV-2 detection in nasopharyngeal swab and saliva. J Infect. 2020 Aug;81(2):e145-e147. doi: 10.1016/j.jinf.2020.05.071. | Letter to the editor |
| 37 | Jeong HW, Kim SM, Kim HS, Kim YI, Kim JH, Cho JY, Kim SH, Kang H, Kim SG, Park SJ, Kim EH, Choi YK. Viable SARS-CoV-2 in various specimens from COVID-19 patients. Clin Microbiol Infect. 2020 Nov;26(11):1520-1524. doi: 10.1016/j.cmi.2020.07.020. | Case report |
| 38 | Jones DL, Baluja MQ, Graham DW, Corbishley A, McDonald JE, Malham SK, Hillary LS, Connor TR, Gaze WH, Moura IB, Wilcox MH, Farkas K. Shedding of SARS-CoV-2 in feces and urine and its potential role in person-to-person transmission and the environment-based spread of COVID-19. Sci Total Environ. 2020 Dec 20;749:141364. doi: 10.1016/j.scitotenv.2020.141364. | Review |
| 39 | Kam KQ, Yung CF, Maiwald M, Chong CY, Soong HY, Loo LH, Tan NWH, Li J, Nadua KD, Thoon KC. Clinical Utility of Buccal Swabs for Severe Acute Respiratory Syndrome Coronavirus 2 Detection in Coronavirus Disease 2019-Infected Children. J Pediatric Infect Dis Soc. 2020 Jul 13;9(3):370-372. doi: 10.1093/jpids/piaa068. | Missing information |
| 40 | Mehwash Kashif, Sana Iqbal, Muhammad Abdullah KamranWhether Human Saliva a Useful Tool for the Diagnosis of COVID-19, J Res Med Dent Sci, 2020, 8(5): 74-76 | Commentary |
| 41 | Khoury NC, Russi TJ. A case of gastrointestinal-predominant COVID-19 demonstrates value of stool PCR test. J Med Virol. 2020 Aug 20:10.1002/jmv.26448. doi: 10.1002/jmv.26448. | Letter to the editor |
| 42 | Kim JM, Kim HM, Lee EJ, Jo HJ, Yoon Y, Lee NJ, Son J, Lee YJ, Kim MS, Lee YP, Chae SJ, Park KR, Cho SR, Park S, Kim SJ, Wang E, Woo S, Lim A, Park SJ, Jang J, Chung YS, Chin BS, Lee JS, Lim D, Han MG, Yoo CK. Detection and Isolation of SARS-CoV-2 in Serum, Urine, and Stool Specimens of COVID-19 Patients from the R | Missing information |
| 43 | Kipkorir V, Cheruiyot I, Ngure B, Misiani M, Munguti J. Prolonged SARS-CoV-2 RNA detection in anal/rectal swabs and stool specimens in COVID-19 patients after negative conversion in nasopharyngeal RT-PCR test. J Med Virol. 2020 Nov;92(11):2328-2331. doi: 10.1002/jmv.26007. | Letter to the editor |
| 44 | Kunz Y, Horninger W, Pinggera GM. Are urologists in trouble with SARS-CoV-2? Reflections and recommendations for specific interventions. BJU Int. 2020 Dec;126(6):670-678. doi: 10.1111/bju.15141. | Review |
| 45 | Lescure FX, Bouadma L, Nguyen D, Parisey M, Wicky PH, Behillil S, Gaymard A, Bouscambert-Duchamp M, Donati F, Le Hingrat Q, Enouf V, Houhou-Fidouh N, Valette M, Mailles A, Lucet JC, Mentre F, Duval X, Descamps D, Malvy D, Timsit JF, Lina B, van-der-Werf S, Yazdanpanah Y. Clinical and virological data of the first cases of COVID-19 in Europe: a case series. Lancet Infect Dis. 2020 Jun;20(6):697-706. doi: 10.1016/S1473-3099(20)30200-0. | One single samples |
| 46 | Li J, Feng J, Liu TH, Xu FC, Song GQ. An infant with a mild SARS-CoV-2 infection detected only by anal swabs: a case report. Braz J Infect Dis. 2020 May-Jun;24(3):247-249. doi: 10.1016/j.bjid.2020.04.009. | Case report |
| 47 | Liu J, Xiao Y, Shen Y, Shi C, Chen Y, Shi P, Gao Y, Wang Y, Lu B. Detection of SARS-CoV-2 by RT-PCR in anal from patients who have recovered from coronavirus disease 2019. J Med Virol. 2020 Oct;92(10):1769-1771. doi: 10.1002/jmv.25875. | Letter to the editor |
| 48 | Liu R, Han H, Liu F, Lv Z, Wu K, Liu Y, Feng Y, Zhu C. Positive rate of RT-PCR detection of SARS-CoV-2 infection in 4880 cases from one hospital in Wuhan, China, from Jan to Feb 2020. Clin Chim Acta. 2020 Jun;505:172-175. doi: 10.1016/j.cca.2020.03.009. | No information regardinbg TP, FN, TN and FP |
| 49 | Lübke N, Senff T, Scherger S, Hauka S, Andrée M, Adams O, Timm J, Walker A. Extraction-free SARS-CoV-2 detection by rapid RT-qPCR universal for all primary respiratory materials. J Clin Virol. 2020 Sep;130:104579. doi: 10.1016/j.jcv.2020.104579. | No information regardinbg TP, FN, TN and FP |
| 50 | Ma X, Su L, Zhang Y, Zhang X, Gai Z, Zhang Z. Do children need a longer time to shed SARS-CoV-2 in stool than adults? J Microbiol Immunol Infect. 2020 Jun;53(3):373-376. doi: 10.1016/j.jmii.2020.03.010. | Lack of negative cases |
| 51 | Martinez RM. Clinical Samples for SARS-CoV-2 Detection: Review of the Early Literature. Clin Microbiol Newsl. 2020 Aug 1;42(15):121-127. doi: 10.1016/j.clinmicnews.2020.07.001. | Review |
| 52 | McCormick-Baw C, Morgan K, Gaffney D, Cazares Y, Jaworski K, Byrd A, Molberg K, Cavuoti D. Saliva as an Alternate Specimen Source for Detection of SARS-CoV-2 in Symptomatic Patients Using Cepheid Xpert Xpress SARS-CoV-2. J Clin Microbiol. 2020 Jul 23;58(8):e01109-20. doi: 10.1128/JCM.01109-20. | Letter to the editor |
| 53 | Morone G, Palomba A, Iosa M, Caporaso T, De Angelis D, Venturiero V, Savo A, Coiro P, Carbone D, Gimigliano F, Iolascon G, Paolucci S. Incidence and Persistence of Viral Shedding in COVID-19 Post-acute Patients With Negativized Pharyngeal Swab: A Systematic Review. Front Med (Lausanne). 2020 Aug 28;7:562. doi: 10.3389/fmed.2020.00562. | Review |
| 54 | Nagura-Ikeda M, Imai K, Tabata S, Miyoshi K, Murahara N, Mizuno T, Horiuchi M, Kato K, Imoto Y, Iwata M, Mimura S, Ito T, Tamura K, Kato Y. Clinical Evaluation of Self-Collected Saliva by Quantitative Reverse Transcription-PCR (RT-qPCR), Direct RT-qPCR, Reverse Transcription-Loop-Mediated Isothermal Amplification, and a Rapid Antigen Test To Diagnose COVID-19. J Clin Microbiol. 2020 Aug 24;58(9):e01438-20. doi: 10.1128/JCM.01438-20. | Comparison between different methods |
| 55 | Opota O, Brouillet R, Greub G, Jaton K. Comparison of SARS-CoV-2 RT-PCR on a high-throughput molecular diagnostic platform and the cobas SARS-CoV-2 test for the diagnostic of COVID-19 on various clinical samples. Pathog Dis. 2020 Nov 11;78(8):ftaa061. doi: 10.1093/femspd/ftaa061. | No information regardinbg TP, FN, TN and FP |
| 56 | Opota O, Brouillet R, Greub G, Jaton K. Comparison of SARS-CoV-2 RT-PCR on a high-throughput molecular diagnostic platform and the cobas SARS-CoV-2 test for the diagnostic of COVID-19 on various clinical samples. Pathog Dis. 2020 Nov 11;78(8):ftaa061. doi: 10.1093/femspd/ftaa061. | Comparison between different methods |
| 57 | Ozma MA, Maroufi P, Khodadadi E, Köse Ş, Esposito I, Ganbarov K, Dao S, Esposito S, Dal T, Zeinalzadeh E, Kafil HS. Clinical manifestation, diagnosis, prevention and control of SARS-CoV-2 (COVID-19) during the outbreak period. Infez Med. 2020 Ahead of print Jun 1;28(2):153-165. | Review |
| 58 | Ozturker ZK. Conjunctivitis as sole symptom of COVID-19: A case report and review of literature. Eur J Ophthalmol. 2020 Jul 24:1120672120946287. doi: 10.1177/1120672120946287. | Case report |
| 59 | Pan Y, Long L, Zhang D, Yuan T, Cui S, Yang P, Wang Q, Ren S. Potential False-Negative Nucleic Acid Testing Results for Severe Acute Respiratory Syndrome Coronavirus 2 from Thermal Inactivation of Samples with Low Viral Loads. Clin Chem. 2020 Jun 1;66(6):794-801. doi: 10.1093/clinchem/hvaa091. | Comparison between different methods |
| 60 | Park SK, Lee CW, Park DI, Woo HY, Cheong HS, Shin HC, Ahn K, Kwon MJ, Joo EJ. Detection of SARS-CoV-2 in Fecal Samples From Patients With Asymptomatic and Mild COVID-19 in Korea. Clin Gastroenterol Hepatol. 2020 Jun 10:S1542-3565(20)30777-1. doi: 10.1016/j.cgh.2020.06.005. | Only fecal samples |
| 61 | Petrovan V, Vrajmasu V, Bucur AC, Soare DS, Radu E, Dimon P, Zaulet M. Evaluation of Commercial qPCR Kits for Detection of SARS-CoV-2 in Pooled Samples. Diagnostics (Basel). 2020 Jul 11;10(7):472. doi: 10.3390/diagnostics10070472. | Comparison between different methods |
| 62 | Prajapati S, Sharma M, Kumar A, Gupta P, Narasimha Kumar GV. An update on novel COVID-19 pandemic: a battle between humans and virus. Eur Rev Med Pharmacol Sci. 2020 May;24(10):5819-5829. doi: 10.26355/eurrev_202005_21377. | Review |
| 63 | Ren JG, Li DY, Wang CF, Wu JH, Wang Y, Sun YJ, Zhang Q, Wang YY, Chang XJ. Positive RT-PCR in urine from an asymptomatic patient with novel coronavirus 2019 infection: a case report. Infect Dis (Lond). 2020 Aug;52(8):571-574. doi: 10.1080/23744235.2020.1766105. | Letter to the editor |
| 64 | Rawlings SA, Ignacio C, Porrachia M, Du P, Smith DM, Chaillon A. No Evidence of SARS-CoV-2 Seminal Shedding Despite SARS-CoV-2 Persistence in the Upper Respiratory Tract. Open Forum Infect Dis. 2020 Aug 7;7(8):ofaa325. doi: 10.1093/ofid/ofaa325. | Missing information |
| 65 | Ren JG, Li DY, Wang CF, Wu JH, Wang Y, Sun YJ, Zhang Q, Wang YY, Chang XJ. Positive RT-PCR in urine from an asymptomatic patient with novel coronavirus 2019 infection: a case report. Infect Dis (Lond). 2020 Aug;52(8):571-574. doi: 10.1080/23744235.2020.1766105. | Case report |
| 66 | Riccò M, Ranzieri S, Peruzzi S, Valente M, Marchesi F, Balzarini F, Bragazzi NL, Signorelli C. RT-qPCR assays based on saliva rather than on nasopharyngeal swabs are possible but should be interpreted with caution: results from a systematic review and meta-analysis. Acta Biomed. 2020 Sep 7;91(3):e2020025. doi: 10.23750/abm.v91i3.10020. | Review |
| 67 | Ruggiero A, Sanguinetti M, Gatto A, Attinà G, Chiaretti A. Diagnosis of COVID-19 infection in children: Less nasopharyngeal swabs, more saliva. Acta Paediatr. 2020 Sep;109(9):1913-1914. doi: 10.1111/apa.15397. | Letter to the editor |
| 68 | Sapkota D, Thapa SB, Hasséus B, Jensen JL. Saliva testing for COVID-19? Br Dent J. 2020 May;228(9):658-659. doi: 10.1038/s41415-020-1594-7. | Commentary |
| 69 | Sarode GS, Sarode SC, Sengupta N, Gadbail AR, Gondivkar S, Sharma NK, Patil S. Clinical status determines the efficacy of salivary and nasopharyngeal samples for detection of SARS-CoV-2. Clin Oral Investig. 2020 Dec;24(12):4661-4662. doi: 10.1007/s00784-020-03630-9. | Letter to the editor |
| 70 | Sarode SC, Sarode GS, Gopalakrishnan D, Patil S. Critical appraisal on salivary diagnostic for COVID-19. Oral Oncol. 2020 Sep;108:104926. doi: 10.1016/j.oraloncology.2020.104926. | Letter to the editor |
| 71 | Shi D, Wu W, Wang Q, Xu K, Xie J, Wu J, Lv L, Sheng J, Guo J, Wang K, Fang D, Li Y, Li L. Clinical Characteristics and Factors Associated With Long-Term Viral Excretion in Patients With Severe Acute Respiratory Syndrome Coronavirus 2 Infection: a Single-Center 28-Day Study. J Infect Dis. 2020 Aug 17;222(6):910-918. doi: 10.1093/infdis/jiaa388. | Risk factors |
| 72 | Singhal T. A Review of Coronavirus Disease-2019 (COVID-19). Indian J Pediatr. 2020 Apr;87(4):281-286. doi: 10.1007/s12098-020-03263-6. | Review |
| 73 | Sirakaya E, Sahiner M, Sirakaya HA. A Patient with Bilateral Conjunctivitis Positive for Sars-Cov-2 RNA in Conjunctival Sample. Cornea. 2020 Jul 2:10.1097/ICO.0000000000002485. doi: 10.1097/ICO.0000000000002485. | Case report |
| 74 | Skolimowska K, Rayment M, Jones R, Madona P, Moore LSP, Randell P. Non-invasive saliva specimens for the diagnosis of COVID-19: caution in mild outpatient cohorts with low prevalence. Clin Microbiol Infect. 2020 Dec;26(12):1711-1713. doi: 10.1016/j.cmi.2020.07.015. | Letter to the editor |
| 75 | SoRelle JA, Mahimainathan L, McCormick-Baw C, Cavuoti D, Lee F, Thomas A, Sarode R, Clark AE, Muthukumar A. Saliva for use with a point of care assay for the rapid diagnosis of COVID-19. Clin Chim Acta. 2020 Nov;510:685-686. doi: 10.1016/j.cca.2020.09.001. | Letter to the editor |
| 76 | Sri Santosh T, Parmar R, Anand H, Srikanth K, Saritha M. A Review of Salivary Diagnostics and Its Potential Implication in Detection of Covid-19. Cureus. 2020 Apr 17;12(4):e7708. doi: 10.7759/cureus.7708. | Review |
| 77 | Sun CB, Wang YY, Liu GH, Liu Z. Role of the Eye in Transmitting Human Coronavirus: What We Know and What We Do Not Know. Front Public Health. 2020 Apr 24;8:155. doi: 10.3389/fpubh.2020.00155. | Review |
| 78 | Sun D, Zhu F, Wang C, Wu J, Liu J, Chen X, Liu Z, Wu Z, Lu X, Ma J, Peng H, Xiao H. Children Infected With SARS-CoV-2 From Family Clusters. Front Pediatr. 2020 Jun 23;8:386. doi: 10.3389/fped.2020.00386. | One single samples |
| 79 | Sun J, Xiao J, Sun R, Tang X, Liang C, Lin H, Zeng L, Hu J, Yuan R, Zhou P, Peng J, Xiong Q, Cui F, Liu Z, Lu J, Tian J, Ma W, Ke C. Prolonged Persistence of SARS-CoV-2 RNA in Body Fluids. Emerg Infect Dis. 2020 Aug;26(8):1834-1838. doi: 10.3201/eid2608.201097. | Comparison between symptomatic and asymptomatic patients |
| 80 | Sun J, Zhu A, Li H, Zheng K, Zhuang Z, Chen Z, Shi Y, Zhang Z, Chen SB, Liu X, Dai J, Li X, Huang S, Huang X, Luo L, Wen L, Zhuo J, Li Y, Wang Y, Zhang L, Zhang Y, Li F, Feng L, Chen X, Zhong N, Yang Z, Huang J, Zhao J, Li YM. Isolation of infectious SARS-CoV-2 from urine of a COVID-19 patient. Emerg Microbes Infect. 2020 Dec;9(1):991-993. doi: 10.1080/22221751.2020.1760144. | Letter to the editor |
| 81 | Szymczak WA, Goldstein DY, Orner EP, Fecher RA, Yokoda RT, Skalina KA, Narlieva M, Gendlina I, Fox AS. Utility of Stool PCR for the Diagnosis of COVID-19: Comparison of Two Commercial Platforms. J Clin Microbiol. 2020 Aug 24;58(9):e01369-20. doi: 10.1128/JCM.01369-20. | Comparison between different methods |
| 82 | Tajima Y, Suda Y, Yano K. A case report of SARS-CoV-2 confirmed in saliva specimens up to 37 days after onset: Proposal of saliva specimens for COVID-19 diagnosis and virus monitoring. J Infect Chemother. 2020 Oct;26(10):1086-1089. doi: 10.1016/j.jiac.2020.06.011. | Case report |
| 83 | Takeuchi Y, Furuchi M, Kamimoto A, Honda K, Matsumura H, Kobayashi R. Saliva-based PCR tests for SARS-CoV-2 detection. J Oral Sci. 2020;62(3):350-351. doi: 10.2334/josnusd.20-0267. | Commentary |
| 84 | Liu J, Xiao Y, Shen Y, Shi C, Chen Y, Shi P, Gao Y, Wang Y, Lu B. Detection of SARS-CoV-2 by RT-PCR in anal from patients who have recovered from coronavirus disease 2019. J Med Virol. 2020 Oct;92(10):1769-1771. doi: 10.1002/jmv.25875. | Letter to the editor |
| 85 | To KK, Tsang OT, Leung WS, Tam AR, Wu TC, Lung DC, Yip CC, Cai JP, Chan JM, Chik TS, Lau DP, Choi CY, Chen LL, Chan WM, Chan KH, Ip JD, Ng AC, Poon RW, Luo CT, Cheng VC, Chan JF, Hung IF, Chen Z, Chen H, Yuen KY. Temporal profiles of viral load in posterior oropharyngeal saliva samples and serum antibody responses during infection by SARS-CoV-2: an observational cohort study. Lancet Infect Dis. 2020 May;20(5):565-574. doi: 10.1016/S1473-3099(20)30196-1. | Only oropharyngeal samples |
| 86 | To KK, Tsang OT, Yip CC, Chan KH, Wu TC, Chan JM, Leung WS, Chik TS, Choi CY, Kandamby DH, Lung DC, Tam AR, Poon RW, Fung AY, Hung IF, Cheng VC, Chan JF, Yuen KY. Consistent Detection of 2019 Novel Coronavirus in Saliva. Clin Infect Dis. 2020 Jul 28;71(15):841-843. doi: 10.1093/cid/ciaa149. | Only saliva samples |
| 87 | Torretta S, Zuccotti G, Cristofaro V, Ettori J, Solimeno L, Battilocchi L, D'Onghia A, Bonsembiante A, Pignataro L, Marchisio P, Capaccio P. Diagnosis of SARS-CoV-2 by RT-PCR Using Different Sample Sources: Review of the Literature. Ear Nose Throat J. 2020 Aug 31:145561320953231. doi: 10.1177/0145561320953231. | Review |
| 88 | Walsh KA, Jordan K, Clyne B, Rohde D, Drummond L, Byrne P, Ahern S, Carty PG, O'Brien KK, O'Murchu E, O'Neill M, Smith SM, Ryan M, Harrington P. SARS-CoV-2 detection, viral load and infectivity over the course of an infection. J Infect. 2020 Sep;81(3):357-371. doi: 10.1016/j.jinf.2020.06.067. | Review |
| 89 | Wang W, Xu Y, Gao R, Lu R, Han K, Wu G, Tan W. Detection of SARS-CoV-2 in Different Types of Clinical Specimens. JAMA. 2020 May 12;323(18):1843-1844. doi: 10.1001/jama.2020.3786. | Letter to the editor |
| 90 | Williams E, Bond K, Zhang B, Putland M, Williamson DA. Saliva as a Noninvasive Specimen for Detection of SARS-CoV-2. J Clin Microbiol. 2020 Jul 23;58(8):e00776-20. doi: 10.1128/JCM.00776-20. | Letter to the editor |
| 91 | Wong MC, Huang J, Lai C, Ng R, Chan FKL, Chan PKS. Detection of SARS-CoV-2 RNA in fecal specimens of patients with confirmed COVID-19: A meta-analysis. J Infect. 2020 Aug;81(2):e31-e38. doi: 10.1016/j.jinf.2020.06.012. | Review |
| 92 | Wong RC, Wong AH, Ho YI, Leung EC, Lai RW. Performance evaluation of Panther Fusion SARS-CoV-2 assay for detection of SARS-CoV-2 from deep throat saliva, nasopharyngeal, and lower-respiratory-tract specimens. J Med Virol. 2020 Sep 30:10.1002/jmv.26574. doi: 10.1002/jmv.26574. | Comparison between different methods |
| 93 | Wong RC, Wong AH, Ho YI, Leung EC, Lai RW. Evaluation on testing of deep throat saliva and lower respiratory tract specimens with Xpert Xpress SARS-CoV-2 assay. J Clin Virol. 2020 Oct;131:104593. doi: 10.1016/j.jcv.2020.104593. | Comparison between different methods |
| 94 | Wu J, Liu J, Li S, Peng Z, Xiao Z, Wang X, Yan R, Luo J. Detection and analysis of nucleic acid in various biological samples of COVID-19 patients. Travel Med Infect Dis. 2020 Sep-Oct;37:101673. doi: 10.1016/j.tmaid.2020.101673. | Letter to the editor |
| 95 | Wu Y, Liu C, Dong L, Zhang C, Chen Y, Liu J, Zhang C, Duan C, Zhang H, Mol BW, Dennis CL, Yin T, Yang J, Huang H. Coronavirus disease 2019 among pregnant Chinese women: case series data on the safety of vaginal birth and breastfeeding. BJOG. 2020 Aug;127(9):1109-1115. doi: 10.1111/1471-0528.16276. | Case series |
| 96 | Wyllie AL, Fournier J, Casanovas-Massana A, Campbell M, Tokuyama M, Vijayakumar P, Warren JL, Geng B, Muenker MC, Moore AJ, Vogels CBF, Petrone ME, Ott IM, Lu P, Venkataraman A, Lu-Culligan A, Klein J, Earnest R, Simonov M, Datta R, Handoko R, Naushad N, Sewanan LR, Valdez J, White EB, Lapidus S, Kalinich CC, Jiang X, Kim DJ, Kudo E, Linehan M, Mao T, Moriyama M, Oh JE, Park A, Silva J, Song E, Takahashi T, Taura M, Weizman OE, Wong P, Yang Y, Bermejo S, Odio CD, Omer SB, Dela Cruz CS, Farhadian S, Martinello RA, Iwasaki A, Grubaugh ND, Ko AI. Saliva or Nasopharyngeal Swab Specimens for Detection of SARS-CoV-2. N Engl J Med. 2020 Sep 24;383(13):1283-1286. doi: 10.1056/NEJMc2016359. | Commentary |
| 97 | Li X, Chan JF, Li KK, Tso EY, Yip CC, Sridhar S, Chung TW, Chiu KH, Hung DL, Wu AK, Chau SK, Liu R, Lung KC, Tam AR, Cheng VC, To KK, Chan KH, Hung IF, Yuen KY. Detection of SARS-CoV-2 in conjunctival secretions from patients without ocular symptoms. Infection. 2020 Sep 17:1–9. doi: 10.1007/s15010-020-01524-2. | Case series |
| 98 | Xiao F, Sun J, Xu Y, Li F, Huang X, Li H, Zhao J, Huang J, Zhao J. Infectious SARS-CoV-2 in Feces of Patient with Severe COVID-19. Emerg Infect Dis. 2020 Aug;26(8):1920-1922. doi: 10.3201/eid2608.200681. | Letter to the editor |
| 99 | Xie C, Jiang L, Huang G, Pu H, Gong B, Lin H, Ma S, Chen X, Long B, Si G, Yu H, Jiang L, Yang X, Shi Y, Yang Z. Comparison of different samples for 2019 novel coronavirus detection by nucleic acid amplification tests. Int J Infect Dis. 2020 Apr;93:264-267. doi: 10.1016/j.ijid.2020.02.050. | Letter to the editor |
| 100 | Xing Y, Ni W, Wu Q, Li W, Li G, Wang W, Tong J, Song X, Wong GWK, Xing Q. Dynamics of faecal SARS-CoV-2 in infected children during the convalescent phase. J Infect. 2020 Aug;81(2):318-356. doi: 10.1016/j.jinf.2020.03.049. | Letter to the editor |
| 101 | Xu T, Wang J, Hu B, Zhang G, Zhou W, Zheng M, Shen B, Sun B, Zhang Y, Chen Y, Yu J, Liang M, Pan J, Chen C, Chen H, Jiang M, Xu L, Qu J, Chen JF. Identification of the RNase-binding site of SARS-CoV-2 RNA for anchor primer-PCR detection of viral loading in 306 COVID-19 patients. Brief Bioinform. 2020 Sep 16:bbaa193. doi: 10.1093/bib/bbaa193. | One single samples |
| 102 | Xu Y, Li X, Zhu B, Liang H, Fang C, Gong Y, Guo Q, Sun X, Zhao D, Shen J, Zhang H, Liu H, Xia H, Tang J, Zhang K, Gong S. Characteristics of pediatric SARS-CoV-2 infection and potential evidence for persistent fecal viral shedding. Nat Med. 2020 Apr;26(4):502-505. doi: 10.1038/s41591-020-0817-4. | Letter to the editor |
| 103 | Xu Y, Cheng M, Chen X, Zhu J. Current approaches in laboratory testing for SARS-CoV-2. Int J Infect Dis. 2020 Nov;100:7-9. doi: 10.1016/j.ijid.2020.08.041. | Letter to the editor |
| 104 | Tong Y, Bao A, Chen H, Huang J, Lv Z, Feng L, Cheng Y, Wang Y, Bai L, Rao W, Zheng H, Wu Z, Qiao B, Zhao Z, Wang H, Li Y. Necessity for detection of SARS-CoV-2 RNA in multiple types of specimens for the discharge of the patients with COVID-19. J Transl Med. 2020 Nov 2;18(1):411. doi: 10.1186/s12967-020-02580-w. | No information regardinbg TP, FN, TN and FP |
| 105 | Yee R, Truong T, Pannaraj PS, Eubanks N, Gai E, Jumarang J, Turner L, Peralta A, Lee Y, Dien Bard J. Saliva is a Promising Alternative Specimen for the Detection of SARS-CoV-2 in Children and Adults. J Clin Microbiol. 2020 Nov 25:JCM.02686-20. doi: 10.1128/JCM.02686-20. Epub ahead of print. PMID: 33239380. | Missing information |
| 106 | Yoon JG, Yoon J, Song JY, Yoon SY, Lim CS, Seong H, Noh JY, Cheong HJ, Kim WJ. Clinical Significance of a High SARS-CoV-2 Viral Load in the Saliva. J Korean Med Sci. 2020 May 25;35(20):e195. doi: 10.3346/jkms.2020.35.e195. | Case report |
| 107 | Yu C, Li L, Tuersun Y, Zhao X, Feng Q, Zhang T, Tay FR, Ma J. Oropharyngeal Secretion as Alternative for SARS-CoV-2 Detection. J Dent Res. 2020 Sep;99(10):1199-1205. doi: 10.1177/0022034520940292. Epub 2020 Jul 2. PMID: 32613877. | Only oropharyngeal samples |
| 108 | Yuan Y, Wang N, Ou X. Caution should be exercised for the detection of SARS-CoV-2, especially in the elderly. J Med Virol. 2020 Sep;92(9):1641-1648. doi: 10.1002/jmv.25796. | Missing information |
| 109 | Zhen-Dong Y, Gao-Jun Z, Run-Ming J, Zhi-Sheng L, Zong-Qi D, Xiong X, Guo-Wei S. Clinical and transmission dynamics characteristics of 406 children with coronavirus disease 2019 in China: A review. J Infect. 2020 Aug;81(2):e11-e15. doi: 10.1016/j.jinf.2020.04.030. | Review |
| 110 | Zhu J, Guo J, Xu Y, Chen X. Viral dynamics of SARS-CoV-2 in saliva from infected patients. J Infect. 2020 Sep;81(3):e48-e50. doi: 10.1016/j.jinf.2020.06.059. | Letter to the editor |
| 111 | Zhurakivska K, Troiano G, Pannone G, Caponio VCA, Lo Muzio L. An Overview of the Temporal Shedding of SARS-CoV-2 RNA in Clinical Specimens. Front Public Health. 2020 Aug 20;8:487. doi: 10.3389/fpubh.2020.00487. | Review |
| 112 | Zhou Y, Duan C, Zeng Y, Tong Y, Nie Y, Yang Y, Chen Z, Chen C. Ocular Findings and Proportion with Conjunctival SARS-COV-2 in COVID-19 Patients. Ophthalmology. 2020 Jul;127(7):982-983. doi: 10.1016/j.ophtha.2020.04.028. | Commentary |

### **Appendix S3.** Methodological quality of the included studies (QUADAS-2).

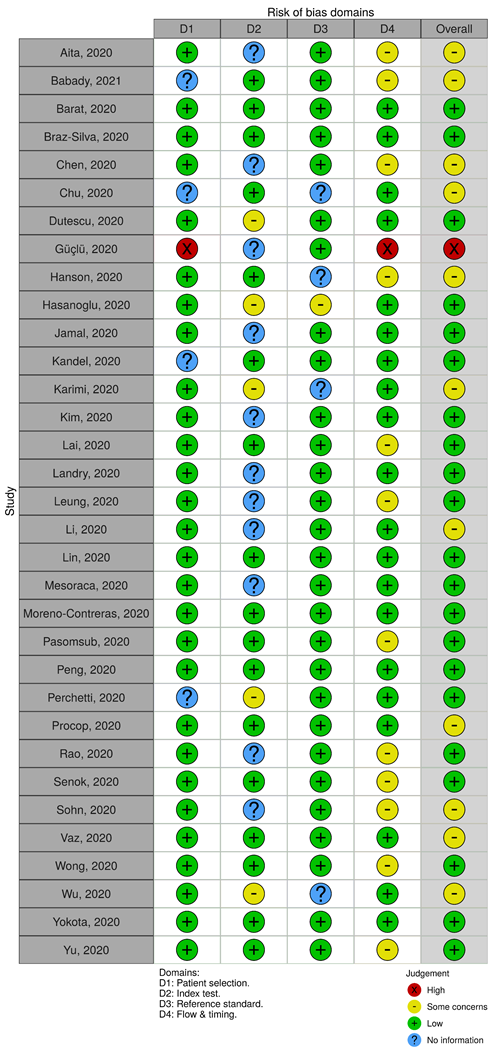

### **Appendix S4.** Meta-regressions.

|  | **Sensitivity** | | **Specificity** | |
| --- | --- | --- | --- | --- |
| **Index Specimen** | **Coefficient (SE)** | **p-value** | **Coefficient (SE)** | **p-value** |
| **Saliva** | | | | |
| M/F Ratio | 0.00 (0.04) | 0.994 | 0.01 (0.02) | 0.441 |
| N | 0.00 (0.00) | 0.518 | **0.00 (0.00)** | **0.034** |
| **DTS/POS** | | | | |
| M/F Ratio | -0.31 (0.21) | 0.132 | **-3.40 (0.99)** | **<0.001** |
| N | -0.00 (0.00) | 0.607 | 0.00 (0.00) | 0.109 |
| **Tears** | | | | |
| M/F Ratio | -0.19 (0.15) | 0.198 | **0.47 (0.23)** | **0.037** |
| N | -0.01 (0.00) | 0.264 | 0.01 (0.01) | 0.085 |
| **Feces** | | | | |
| M/F Ratio | -0.48 (0.36) | 0.184 | -0.99 (1.00) | 0.321 |
| N | -0.07 (0.184) | 0.184 | -0.14 (0.14) | 0.321 |

M/F – Male/Female, SE – Standard Error, DTS/POS – Depp throat saliva/Posterior Oral Saliva

### **Appendix S5.** Subgroup analysis on tears sensitivity.

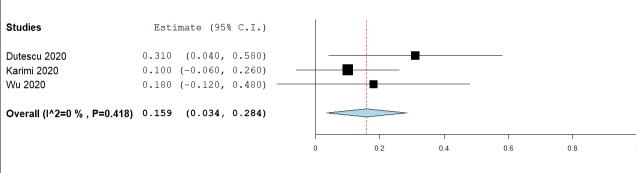

### **Appendix S6.** Subgroup analysis on tears specificity.

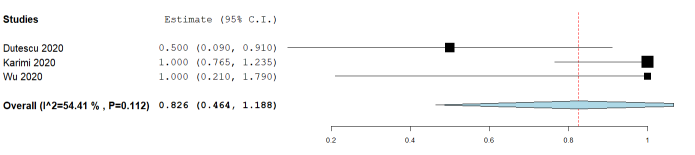

### **Appendix S7.** Subgroup analysis on feces sensitivity.

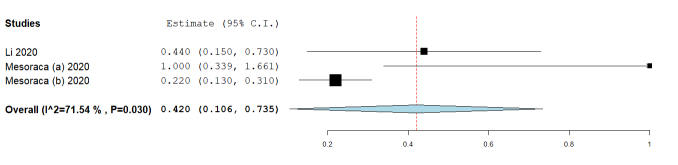

### **Appendix S8.** Subgroup analysis on feces specificity.

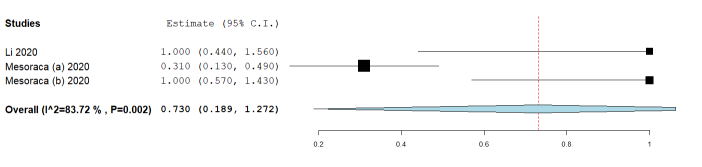

### **Appendix S9.** Forest plots of mean difference to reference sample (NPS) of CT for Saliva. Positive values mean increased RT-PCR cycle (and vice-versa) needed for reliable outcome in reference to NPS.

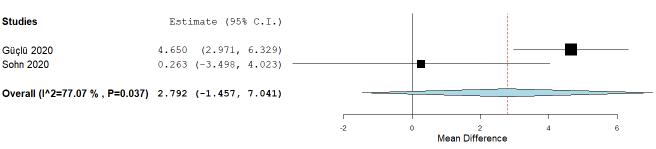

### **Appendix S10.** Forest plots of mean difference to reference sample (NPS) of CT for DTS/POS. Negative values mean decreased RT-PCR cycle (and vice-versa) needed for reliable outcome in reference to NPS.

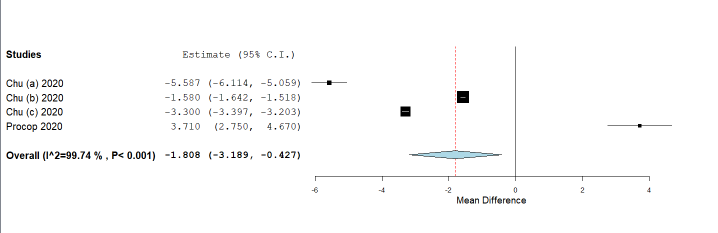
